# Cognition and Future Depression: Associations with Risk in Those With and Without a History of Depression

**DOI:** 10.1101/2025.11.14.25340251

**Authors:** A.N. de Cates, A. Lee, L. Winchester, K.P. Ebmeier, P. Lalousis, R. Upthegrove, S.E. Murphy, C.J. Harmer, T. Nichols, A Topiwala

## Abstract

**Introduction:** Cognitive impairments are common in depression and often persist beyond mood resolution. However, the relationship between cognitive performance, its neurological underpinnings, and future depression risk is unclear, limiting strategies for primary and secondary prevention. Our objective was to determine whether cognition associates with subsequent depression, including both relapse and first-episode occurrences.

**Methods:** 2094 UK Biobank participants with previous ICD-10-defined depression currently in remission (RD) (mean(SD) age: 52.4(7.25) years) were age- and sex-matched to 2094 participants without depression history or current antidepressant use. Cognitive scores were compared between groups at the composite (z-score), domain, and task levels. MRI-derived phenotypes assessed brain network structure and functional connectivity. Longitudinal associations with future depression were assessed using logistic regression models controlling for key confounders.

**Results:** Participants with RD had a higher risk of future depression (30%) than controls (8.5%). Composite cognitive performance in controls was inversely associated with future depression risk (risk estimated marginal means: 0.48% at -1SD, 0.37% at mean, 0.28% at +1SD). In RD, this relationship was reversed (1.56% at -1SD, 1.80% at mean, 2.08% at +1SD). Executive functioning, processing speed, and reasoning task scores all contributed. Higher grey matter in default mode network regions was associated with better concurrent cognitive performance across all participants, but not with future depression risk. Other MRI findings were limited.

**Conclusion:** RD carried a threefold higher risk of future depression than controls. Cognitive performance was a risk marker for future depression in both groups but in opposing directions. Neuroimaging metrics provided little predictive value.

**What is already known on this topic:** Neurocognitive impairments are common in depression, even after low mood has resolved and outside of comorbid neurodegenerative processes. However, the specific relationship between cognitive performance and risk of future depression is unclear, including how this relates to previous history of depression.

**What this study adds:** Cognitive performance is a differential risk marker for future depression in those with previous depression compared to matched controls: without previous history of depression, poorer cognitive scores confer the highest risk; in remitted depression, higher levels of cognitive performance are associated with greater risk of depressive relapse.

**How this study might affect research, practice or policy:** Further research should explore targeting interventions based on specific cognitive profiles, especially in high-risk populations such as those with previous episodes of depression.

## BACKGROUND

Major depressive disorder affects one in five people in their lifetime with significant morbidity and mortality ^1^, and poor cognition is a main component of the diagnosis for all ages, affecting 70-90% of those affected ^2^ ^3^. Individuals with a history of depression appear to face a significantly elevated risk of relapse ^1^, but the ability to predict which individuals are most susceptible remains limited – despite its potential to inform targeted prevention and improve clinical outcomes. Cognitive impairments persist in approximately 40% of patients (including deficits in attention and executive functioning, processing speed, and learning and memory), even after mood symptoms remit ^4–6^ and respond poorly to first-line antidepressants ^7–9^. Cognitive problems also tend to worsen with successive episodes ^4^ ^5^ ^10^, suggesting that they hold promise as a robust prospective marker for depressive relapse. However, despite the potential clinical importance, longitudinal evidence directly linking cognitive deficits to future depression risk is currently lacking, including details of which cognitive domains may be most relevant for relapse and therefore a potential target for preventative measures.

A proposed mechanistic link between poor cognition and depressive relapse is dysconnectivity within the default mode (DMN), central executive (cEN), and salience brain networks (SN) (also known as the “triple network” ^11^). Alterations in the triple network are noted in both current and remitted depression compared to controls ^12–14^, and these networks also directly link to cognitive performance (i.e. worse cognition is associated with weaker DMN-SN and SN-cEN connections) ^15^. This suggests both impaired cognition and network connectivity may mark relapse risk ^16^ ^17^, but uncertainty remains as to how the structural and functional integrity of these networks might interact with cognition to predict future depressive episodes.

## OBJECTIVE

Using UK Biobank data, a large, prospective, multimodal study aimed to identify cognitive and neural markers of depression onset and relapse vulnerability. We also assessed whether structural and functional properties of the triple network mediate these associations. We hypothesised that poorer cognition would predict future depression, particularly in RD participants (primary analyses), and that abnormalities within the triple network and constituent brain regions would contribute to this risk (secondary analyses).

## METHODS

### Study sample

An overview of methodology is shown in Figure 1. The study comprised participants from the UK Biobank as outlined in the Supplementary Material (Supplementary Figure 1; see completed STROBE statement). The UK Biobank is a prospective cohort study involving approximately 500LJ000 participants from the UK. These individuals were aged 40 to 69 years at the time of their initial assessment visit, conducted between 2006 and 2010. Participants provided informed consent via electronic signature at the time of recruitment. Additional details regarding the UK Biobank protocol can be accessed through their website (http://www.ukbiobank.ac.uk/). The ethical approval for UK Biobank has been granted by the National Information Governance Board for Health and Social Care and the NHS North-West Multi-centre Research Ethics Committee.

**Figure 1:**
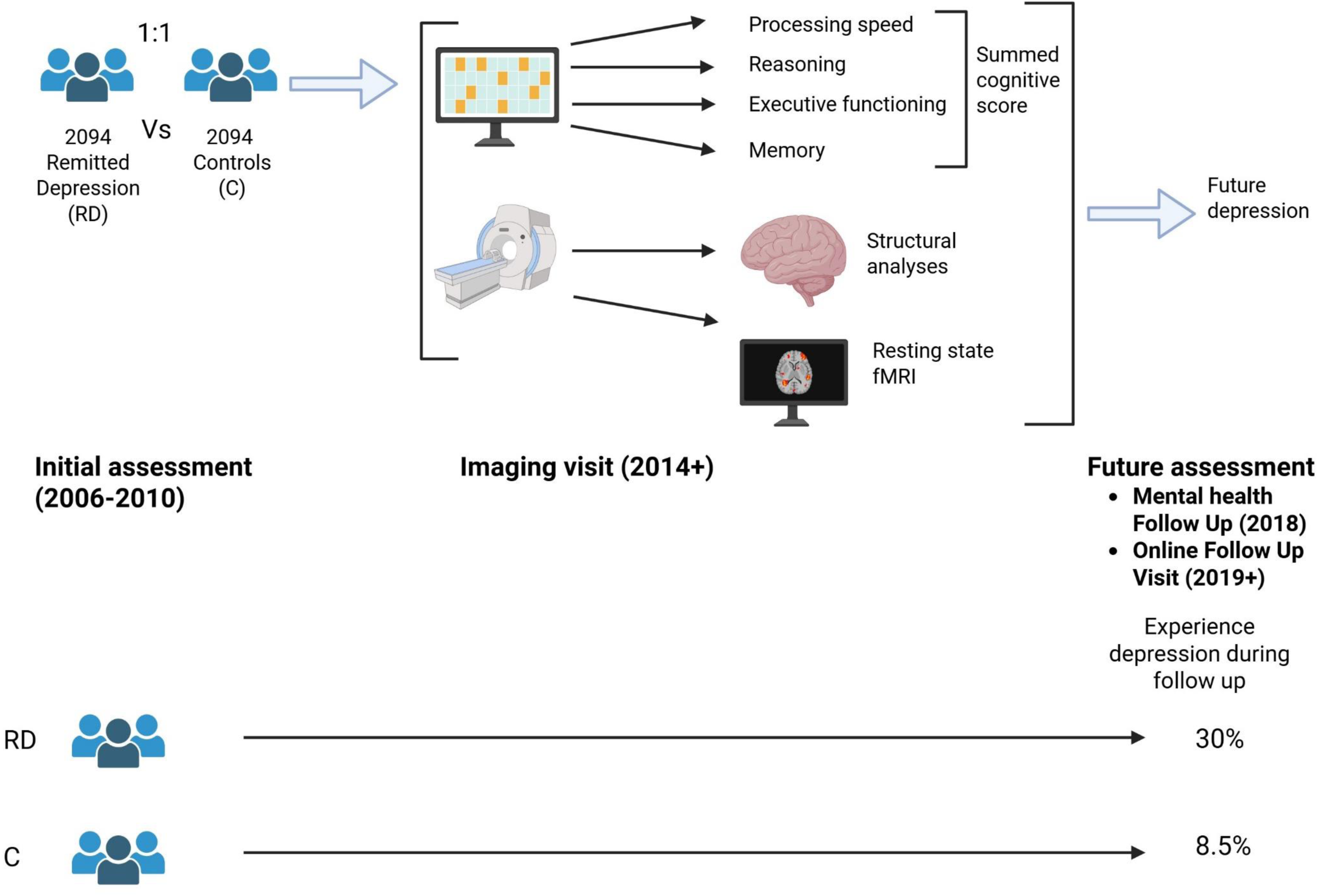
Overview of study methodology.

Sociodemographic data used in this study were from the initial assessment visit (2006–2010) and the first imaging visit (2014–2019). Baseline cognitive measures and MRI data used in the study were collected during the first imaging visit.

Remitted depression was defined as (i) a recorded depressive episode occurring up to 15 years before the first imaging visit, AND (ii) remission from depressive symptoms at the first imaging visit defined as scoring 1 (not at all) or 2 (several days but less than half the days) on each of the two self-report Patient Health Questionnaire (PHQ)-2 core questions (self-reported frequency of low mood and anhedonia symptoms in the last two weeks). For the primary cohort, an ICD-10 diagnosis of at least moderate severity depression using first occurrence and hospital episode statistics (FO+HES) data was used for (i). We also conducted sensitivity analyses altering the threshold for depressive episode using: (a) General Practice (GP) ICD codes for moderate to severe ICD-10 depression for those with linked primary care records, and (b) the “probable depression” UKB-derived algorithm derived from self-report variables. Details of codes used in cohort definitions are available in Supplementary Data.

Remitted depressed cohort participants with confounder, cognitive and fMRI data were matched 1:1 with control participants (with no history of depression) drawn from the remainder of the UK Biobank based on availability of required data, sex and age +/- 2 years, and no lifetime history of ICD-10 coded depression or antidepressant use at the imaging visit (using tabular data antidepressant codes). We also excluded from all RD and control cohorts those with current significant neurological/neurodegenerative disorders, or development of these within the following two years (see Supplementary Methods). Other mental disorders were included as potential confounders.

### MRI acquisition and data processing

The imaging data were obtained on Siemens Skyra 3T MRI scanners equipped with 32-channel head coils. The UK Biobank team performed image processing, quality control checks and automated brain tissue volume computations; their imaging-derived phenotypes (IDPs) were made available to the researchers. The brain imaging protocol used in the UK Biobank includes structural, diffusion and functional imaging from six distinct modalities: T1-weighted, T2-weighted flair, diffusion MRI, susceptibility-weighted imaging, task functional MRI and resting-state functional MRI time series data ^18^.

T1-weighted measures estimated grey matter and cortical measures. Structural regions of the triple network were pre-specified (detailed in Supplementary Table 15(a)). Resting-state functional MRI was conducted at two distinct dimensionalities (25 and 100), resulting in 21 and 55 signal networks, providing information on measures of both within-network and between-network functional connectivity. Additional details on the acquisition parameters, image processing, and specific measurements derived from both imaging modalities are available in Supplementary Methods.

### Details of cognitive assessment

Cognitive behavioural tasks were performed at the imaging visit. Individual tasks were defined by the cognitive domain being tested: (i) Processing speed=Reaction Time [‘Snap-card game’] (UKB_ID_20023:mean time to correctly identify matches from 8 trials); and Symbol-Digit Substitution (UKB_ID_23324:number of symbol digit matches made correctly); (ii) Reasoning=Verbal-Numerical Reasoning [‘Fluid Intelligence’] (UKB_ID_10016:sum of correct answers / fluid intelligence score) and Matrix Pattern Completion (UKB_ID_6373:number of puzzles correctly solved); (iii) Attention and executive function=Digit Span [‘Numeric Memory’] (UKB_ID_4282:maximum digits recalled), Trails A and B (UKB_ID_6350: duration to complete alphanumeric path #2 MINUS 6348:duration to complete numeric path #1), Tower Rearranging (number of puzzles correct); (iv) Memory=Verbal Paired Associates (UKB_ID_20197:number of word pairs correctly associated). The psychometric properties of these tasks have previously been reported ^19^. Calculation of a composite cognitive score was based on previous methodology ^20^. calculating a composite z-score using the mean scores for each task. Separate z-scores were also created for each cognitive domain and each individual task. Participants with incomplete data were excluded from the main composite cognitive analyses. However, if data from at least three tasks were available, missing values were imputed and these participants were included in a sensitivity analysis (see Statistical Analyses). For analyses of individual cognitive domains and tasks, participants were included when data were available.

### Longitudinal outcome data

A depressive episode occurring in either the remitted depressed or matched cohorts after the imaging visit was identified using one of the following: (i) presence of a HES depression code after the imaging time point; OR (ii) a PHQ-9 score greater than 9 on Mental Health Follow Up (MHFU) data (collected in 2018) if imaging visit occurred before MHFU; OR (iii) MHFU age at last episode of depression if greater than age at imaging visit; OR (iv) recent depression recorded at the Online Follow Up visit (collected from 2019).

Models for both primary (cognitive) and secondary (neural/neurocognitive) analyses were pre-specified and are listed in Supplementary Methods (p9). We adjusted for potential confounders, which were self-reported or established at the time of the MRI scan (apart from sex, ethnicity, deprivation score, and education which were assessed at the initial assessment visit). A full list of these is given in the Supplementary Material. In brief, Townsend deprivation is a measure of material deprivation based on address census information. Educational qualifications were recorded as: college or university degree, A level/AS levels or equivalent, O levels/GCSEs or equivalent, CSEs or equivalent, NVQ or HND or HNC or equivalent, other professional qualifications or none of the above. Smoking was reported as: current, previous or never. Alcohol status was as units consumed per week. Body mass index in kg/m^2^. For MRI analyses, age^2^, age^3^, and age-by-sex interaction were accounted for as well as a set of brain imaging-related confounders (see Supplementary Material).

### Statistical analyses

All statistical analysis was performed in RStudio (version 4.4.1). Independent-samples t-tests and chi-square tests were performed to assess potential univariate differences in the sociodemographic characteristics between remitted depressed participants and controls. Binomial logistic regression was performed for both primary (cognitive) and secondary (neural/neurocognitive) analyses, accounting for confounders as detailed above. Where appropriate, regression models were performed with and without group (i.e., remitted depression or control) as a factor to identify relationships across both cohorts. P-values in secondary analyses were corrected for multiple testing using false discovery rate (FDR, 5%).

As well as repeating primary analyses with alternative cohorts as previously described (General Practice (GP) or the “Probable Depression” algorithm data), we performed other sensitivity analyses as follows: i) extending the exclusion of participants who developed neurodegenerative disease or dementia from two to within 10 years after imaging; (ii) excluding participants who had cerebrovascular disease at imaging; (iii) controlling for participants with a close family history of depression. We also repeated analyses after dividing the primary (FO+HES) cohort into participants up to 64 and those over 64 years old. Imputation of missing cognitive data was used as a sensitivity analysis. Prior checks ensured that data were missing at random to confirm the validity of imputation (i.e., individual cognitive task scores, age, and sex were not statistically different in those with and without full cognitive data). The R package MICE was used to impute cognitive data, using predictive mean matching to preserve relationships between variables, and with age and sex included as auxiliary variables using 20 imputations (see Supplementary Material for details). Estimated marginal means were calculated using the emmeans R package.

## FINDINGS

### Sample characteristics

The primary cohort consisted of 2094 participants who met criteria for remitted depression (RD) (Table 1 & Supplementary Figure 1). The mean time interval between record of depressive episode and MRI was 8.15 years (SD 7.70 years). There were incomplete cognitive data for 928 participants from the remitted depression cohort and 877 from the control cohort. Participants with incomplete (n=1805) as opposed to complete (n=2383) cognitive data were not systematically different in terms of age or sex (Age: mean (SD)=52.4 years (7.12 years) complete, mean (SD)=52.8 years (7.31 years) incomplete, t=-1.71, p=0.09; Sex: female:male=59.8:40.2% complete, 58.5:41.5% incomplete, chi-square=0.71, p=0.40). For potentially confounding variables, remitted depressed (RD) participants with incomplete as opposed to complete data comprised a higher proportion of males and people endorsing white ethnicity, and more participants had post-university level education (see Supplementary Table 1(a)).

**Table 1:**
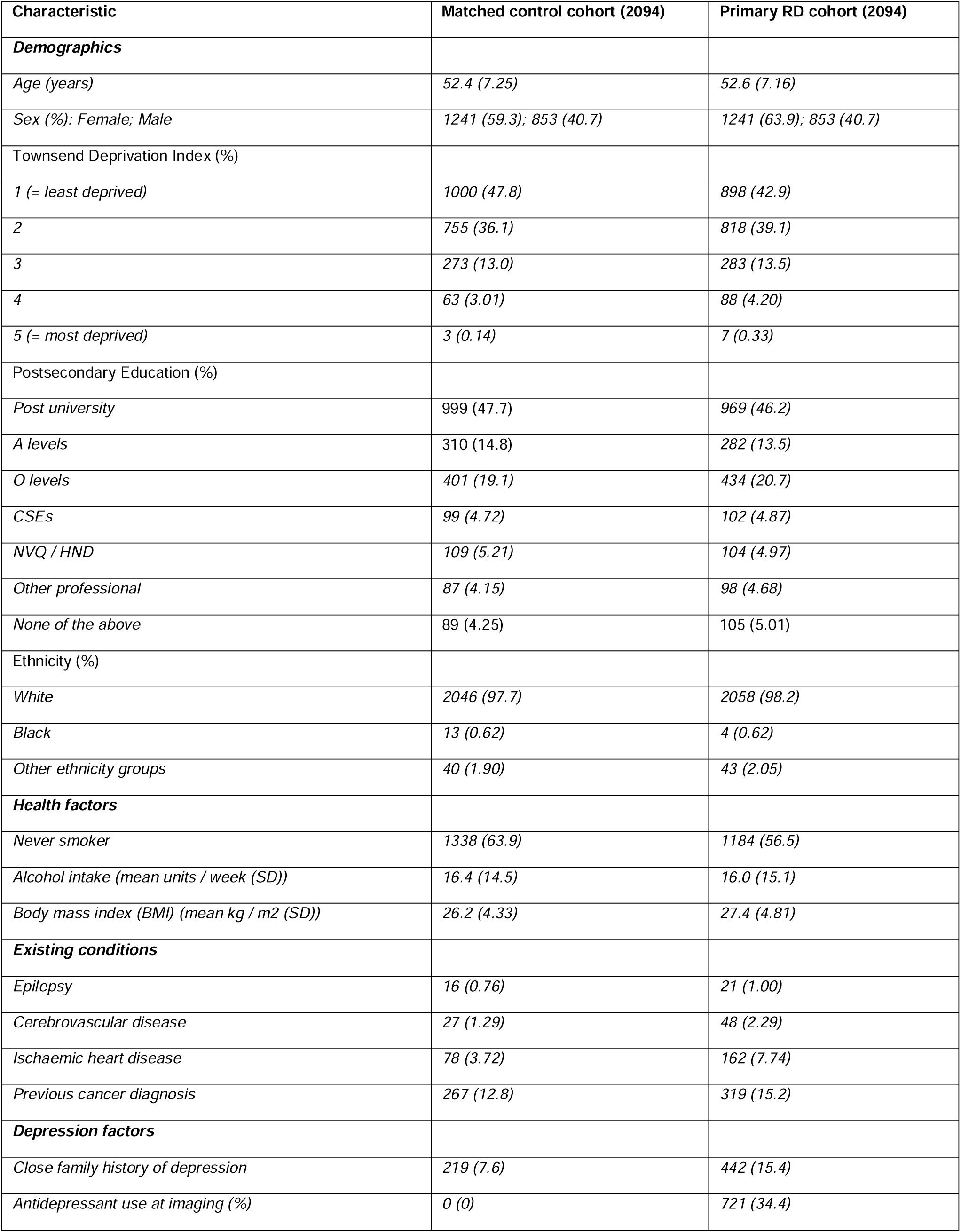
Characteristics and health factors of included participants.

Compared to control participants, the RD group (n=2094) had a significantly higher average BMI, were more likely to be in less affluent socioeconomic groups, and to have pre-existing ischaemic heart disease and a previous diagnosis of cancer at the imaging visit (see Table 1 and Supplementary Table 2(a)). One third of the RD cohort also reported use of antidepressants at this point (34.2%). The RD cohort performed worse on cognitive tasks compared to controls when examined as a composite measure across all domains [F(1,2365)=7.1, p=0.008: composite z-score mean(SD): controls=0.164(0.967); RD=0.065(0.976)]. At the individual domain level, there was a similar pattern with the largest difference in processing speed (see Supplementary Table 3).

Following imaging, 179 (8.5%) controls developed a first episode of depression, while 620 (30%) of the primary RD cohort experienced a relapse of depression (see Supplementary Table 5(b)). Within the RD cohort, 307 individuals developing depression over the follow-up period were receiving antidepressants at the imaging visit (49.5%).

### Cognitive markers of depression onset during follow-up

History of depression (remitted depression versus controls), risk of future depression, and cognitive score at the imaging visit were significantly related [main effect of group: coefficient=0.950, SE=3.29, z=-2.37, p=0.018; composite cognition score*group: coefficient=0.415, SE=0.13, z=3.25, p=0.001 (see Figure 2(A), Supplementary Table 6)]. For control participants, the risk of depression during the follow-up period was low overall, but higher for those with a relatively lower baseline composite cognitive score (mean(SD) depression risk as probability=0.09(0.28), see Figure 2; mean depression risk at mean+/-standard deviation(SD) cognition z-score as estimated marginal means (EMM)=0.0037(0.37%); minus 1SD=0.0048(0.48%), plus 1SD=0.0029(0.29%))). When analysing cognitive domains separately, lower performance on executive functioning, reasoning, and processing speed were all associated with higher depression risk in controls (see Figures 2(C-E), Supplementary Table 7(b-d)). Paradoxically, the opposite pattern of associations was observed in participants with remitted depression – that is, higher composite cognitive performance was associated with greater future depression risk (mean (SD) depression risk as probability=0.30(0.46), see Figure 2; mean depression risk at mean+/-SD cognition z-score as EMM=0.018(1.80%); minus 1SD=0.0156(1.56%), plus 1SD=0.0208(2.08%)). As seen with controls, scores for executive functioning, reasoning, and processing speed all contributed to this finding.

**Figure 2:**
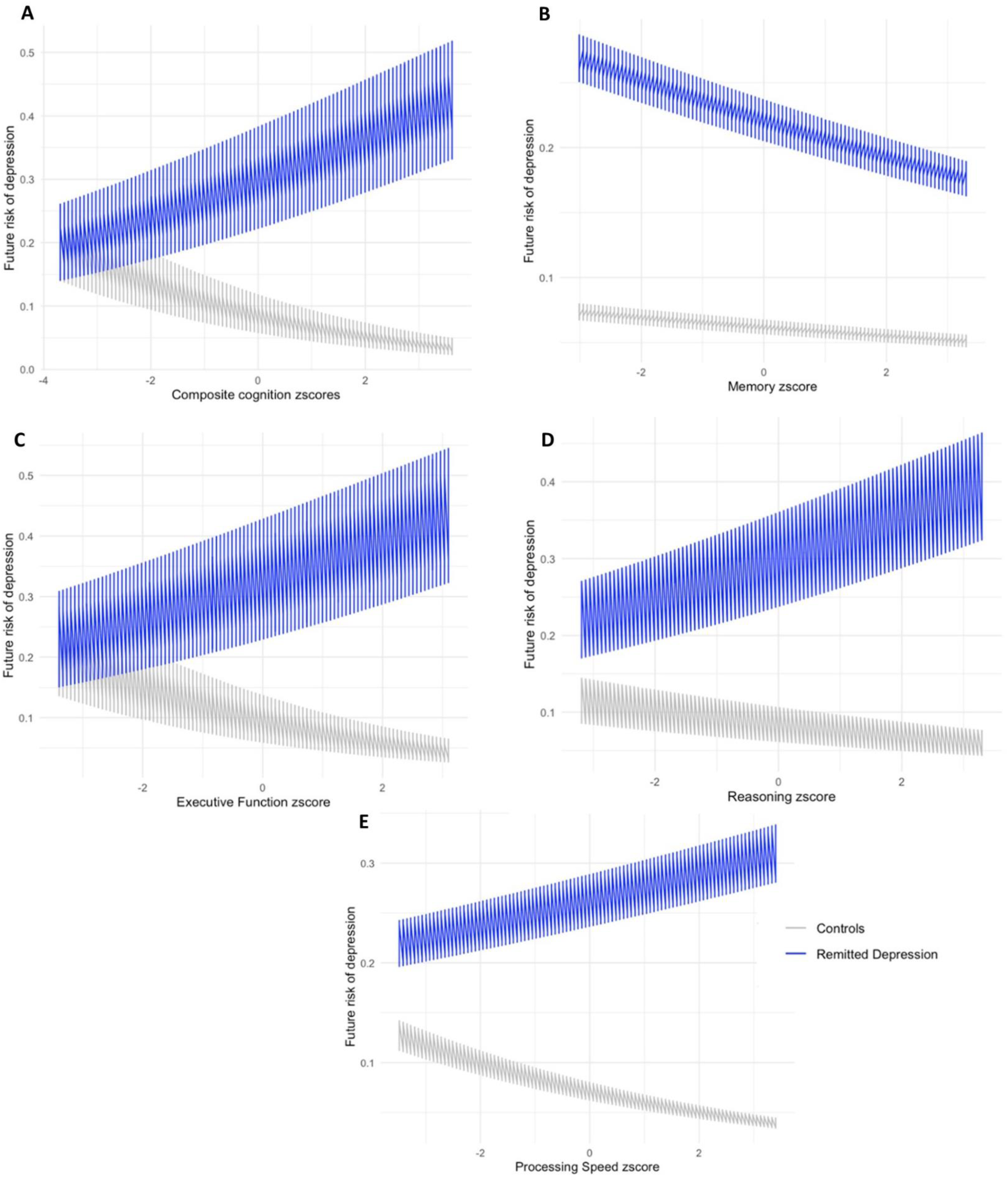
Future depression risk relating to cognition scores at imaging: (A) expressed as composite; (B – E) when considering individual dimensions.

Models without the inclusion of a group factor found no association between composite cognition score and future depression across the two groups [composite cognitive score (coefficient=-0.45, SE=3.18, z=0.102, p=0.918, see Supplementary Table 6 and Supplementary Figure 2; for per domain see Supplementary Table 7(a-d)].

Significant task measure by group interactions for correct digit symbol matches*group (p=0.03), fluid intelligence*group (p=0.03), correct tower arranging*group (p=0.002) appeared to drive the differences seen at the summed dimension and overall composite score level (see Supplementary Table 8 and Supplementary Figure 3).

### Sensitivity analyses

Results for main analyses were similar when imputation was used to account for missing cognitive data (i.e. composite cognitive scores: p=0.03; composite cognitive scores*group: p=0.005, see Supplementary Table 6). Other sensitivity analyses of the primary outcome (association between cognitive performance and future depression) were consistent with the findings from the main analyses (see Supplementary Table 9).

A slightly higher proportion of depressive relapse episodes occurred in the younger subset (64 years and under) of the study population, but there was no significant difference in odds of depression in control and RD groups comparing participants under 65 with those 65 and over (p=0.52; see Supplementary Material p42). Regression analyses using these divided cohorts were limited by reduced power, but results were similar to main analyses, with the composite cognition z-score*group interaction significant in both subgroups (p<0.05, see Supplementary Table 9). Precision (i.e. tightness of confidence intervals) appeared to be greater in the older subset despite the smaller number of participants included (see Supplementary Figure 5).

As post-hoc robustness analyses, we evaluated (i) the effect of antidepressant use on cognitive scores at the imaging visit, (ii) the proportions of participants who overlapped between different RD cohorts depending on definition (first occurrences + hospital episode statistics/General Practice (GP) data/Probable Depression algorithm) and (iii) the demographics between RD and control participants who had a future episode of depression. Antidepressants at baseline and imaging did not appear to have a significant effect on summed cognitive scores at imaging (p > 0.2; see Supplementary Table 10). Proportions of participants who overlapped between primary and other cohorts were similar (see Supplementary Table 11). Demographics were similar between control and RD participants who experienced an episode of depression after the imaging visit (see Supplementary Table 12).

Differences between GP/Probable Depression cohorts and the First Occurrences + Hospital Episode Statistics (FO+HES) primary cohort compared to their respective matched controls are outlined in Supplementary Material. The general pattern of results was similar across both secondary cohorts (i.e. GP records; Probable Depression) compared to the primary cohort (FO+HES) for the primary outcome (association between cognitive performance and future depression). However, results indicated that these secondary cohorts were lower power and less precise with broader confidence intervals when compared to their matched controls cohorts in terms of future risk of depression modelled using both compositive cognitive scores (Supplementary Figure 3) and individual cognitive dimensions (Supplementary Figure 4).

### MRI markers and associations with cognition and future depression

Of the 2094 FO+HES RD cohort used for main analyses, 1554 participants had cognitive data as well as full data for MRI and control variables (see Supplementary Table 1(b) for participant characteristics of this cohort subset). This subset was used for secondary analyses involving MRI data after matching 1:1 to control participants using the same procedure as for primary analyses.

Compared to the control group, the remitted depressed (RD) group had greater head movement and reduced mean grey matter volume. Other imaging parameters were not significantly different between groups (see Supplementary Table 13). There were no significant differences in grey matter volumes of structural regions of triple network (pre-specified) between RD participants when compared at the imaging assessment (see Supplementary Table 15(a) and (b)).

Increased volume of the amygdala, hippocampus, and middle frontal gyrus bilaterally and the left precuneus and left PCC were associated cross-sectionally with better composite cognitive performance across all participants (see Figure 3). These regions are considered part of the default mode network.

**Figure 3:**
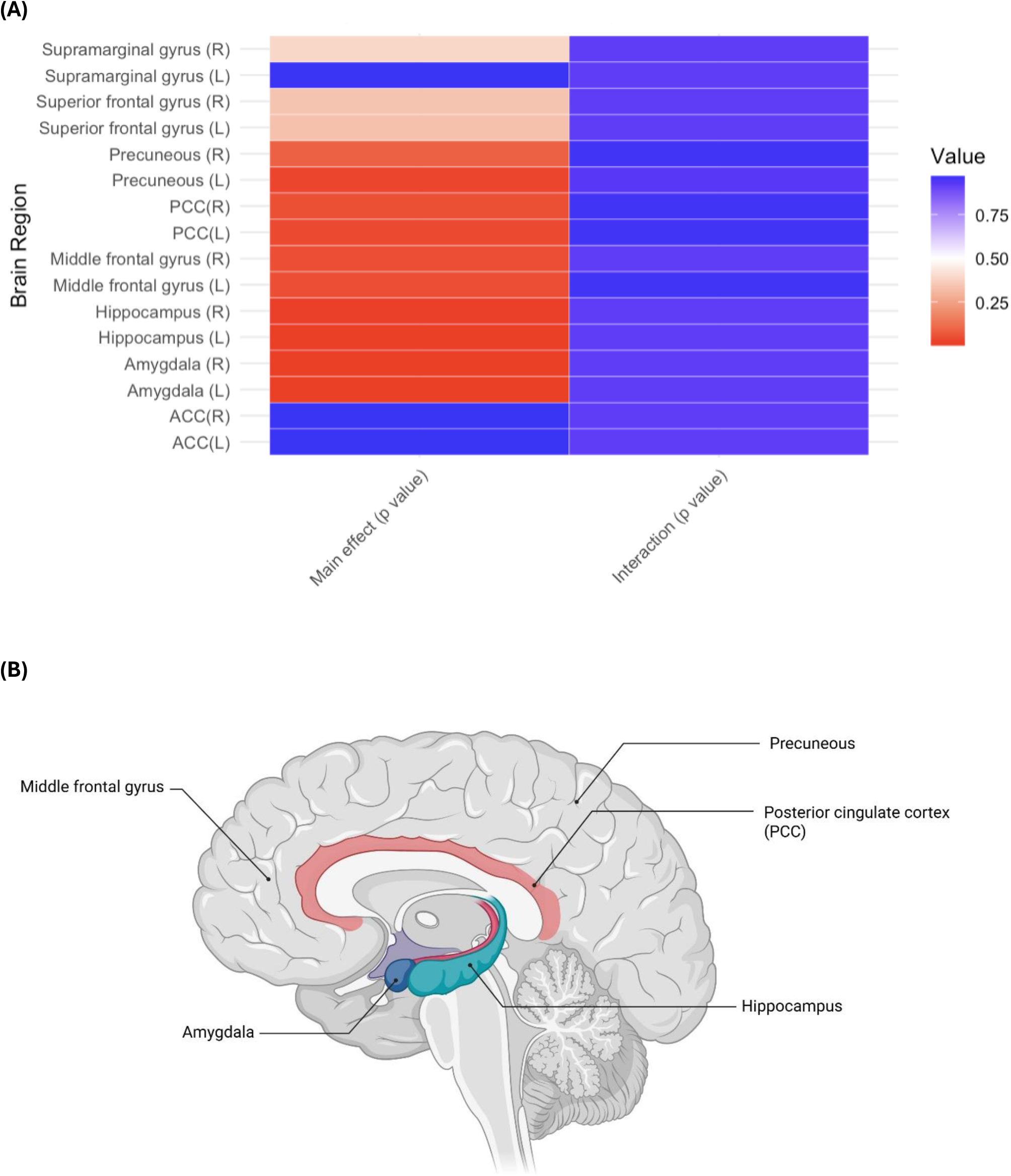
Composite cognition scores and cross-sectional associations with pre-specified triple network regional volumes across all participants (n=1554); (A) Heatmap of significant associations; (B) Mapping of regions equating to default mode network where associations reach significance across all participants.

Structural measures of the triple network were not associated with depression risk amongst control participants or individuals with remitted depression (see Supplementary Table 16). Similar findings were observed for functional connectivity measures of the triple network (see Supplementary Tables 18 and 19).

## DISCUSSION

As hypothesised, lower cognitive performance predicted a higher risk of future depression in participants without a prior history. Those with past depression (RD) showed poorer cognitive function and reduced grey matter volume on average compared to matched controls and were three times more likely than control participants to experience a depressive episode during follow-up. Surprisingly, those RD individuals with the highest cognitive performance appeared at greatest risk of relapse. MRI measures did not explain this vulnerability.

People who have experienced episodes of depression have previously been shown to be at higher risk of further episodes compared to controls, even periods of remission and recovery ^21^. We replicated this finding in our results: 30% of RD participants had a subsequent episode in follow up compared to 8.5% of matched controls. A goal of individualised risk prediction is to identify those within this higher risk RD cohort that are most at risk to enable targeted intervention and potentially improved mental health and functional outcomes. Cognitive functioning has been suggested as a possible risk marker of those at highest risk for relapse in RD ^2^: there is consistent evidence that at least 40% of people with RD perform poorly in terms of cognition across a variety of behavioural tasks ^4^ ^5^, with these impairments worsening with subsequent depressive episodes ^4^ ^5^ ^10^. In this study, we identified that cognitive performance appears to be associated with risk of future depression differently in those with previous depression compared to matched never-depressed controls. That is, lower baseline composite cognitive performance was associated with higher risk of future depression in people who had not experienced depression before, but that relationship was reversed for those with RD (i.e.) higher cognitive performance across a range of tasks was associated with higher future depressive risk. Furthermore, understanding which cognitive domains may be of relevance for longitudinal risk in remitted depression has been relatively under-explored. In our study, consistent with meta-analyses involving remitted depression participants ^22^, we found that cognitive performance on the memory task showed limited (non-significant) impact on future depressive risk between the groups compared to other cognitive dimensions (executive function, processing speed, reaction time).

This study is a novel first step to triangulate cognitive performance and MRI findings with longitudinal depression risk in RD. Our primary analysis findings (i.e. in RD, higher composite cognitive scores were associated with an increased risk of a subsequent depressive episode) was counter to our hypothesis and might seem counterintuitive. This relationship was not evident in control participants, suggesting it was likely not related to the UK Biobank cohort *per se*. However, it is interesting to consider why a difference may exist between RD participants and controls in terms of the relationship between baseline cognitive functioning and future risk of depression. One possible explanation is that adults with RD (who are at greater risk of depression than controls) and with a higher level of cognitive functioning may be more likely than RD participants at a lower cognitive level to notice and report future lower mood symptoms. That is, impaired cognition may increase risk up to a threshold, beyond which people are too impaired to reflect and self-report symptoms (see Supplementary Table 3 for the sample cognitive performance distribution). Alternatively, control participants with no previous depression diagnoses but who subsequently developed depression may have shown a depression prodrome with reduced cognitive functioning in our analyses. Although we excluded participants with pre-existing cardiovascular or neurological disorders or those who developed dementia during the follow-up period, this “late onset depression” may be more aligned with underlying organic causes and thus be more closely associated with worse cognitive performance. A future study assessing cognitive functioning longitudinally alongside evolving risk of future depression may be able to provide more clarity for some of these hypotheses.

The relationship between future relapse risk in RD, abnormal functioning of the triple network, and cognitive performance within RD has been unclear, although both impairments in cognition and the triple network (DMN, cEN, SN) have been suggested as vulnerability markers for future episodes ^16^ ^17^. In RD, functional changes within and between neural networks including the triple network ^13^ ^14^ ^23^ have been found to exist, and in healthy volunteers, connections involving these specific networks also directly link to cognitive performance ^15^. We identified that increased volume of regions of the DMN (amygdala, hippocampus, middle frontal gyrus, precuneus and PCC) were associated with improved cross-sectional cognitive performance across all participants, but this was not specifically associated with a prior history of depression (after controlling for baseline MRI measures). We also demonstrated that compared to the control group, the RD group had reduced grey matter overall. Taken together, this may explain why our previously depressed cohort performed significantly worse on cognitive tasks compared to controls matched for age and sex.

Our analyses had several strengths. The primary RD cohort, using First Occurrences and Hospital Episode Statistics, was carefully constructed with participants that had met ICD-10 diagnostic criteria for depression. We conducted multiple sensitivity, robustness and subgroup analyses to confirm and explore our findings, and imputation to account for any missing cognitive data across participants when calculating the composite cognition score. Sensitivity and robustness analyses indicate that our results are not explained by other factors, such as the presence of cerebrovascular disease, a close family history of depression, age, or the use of antidepressants. Checks completed prior to imputation confirmed that missing cognitive data was missing at random, and analyses with and without imputation were similar, providing reassurance that missing data had minimal impact on results.

When remitted depression was defined using other UK Biobank methodologies (using GP records or using the UK Biobank “Probable Depression” algorithm), results were similar but less precise, supporting findings and suggesting that larger sample sizes are likely to be required if these less stringent definitions are used compared with those based on defining remitted depression using first occurrences and hospital episode statistics. rsfMRI findings may have been affected by limited power due to only a subgroup of participants having full MRI data. Future analyses should also consider including the number of depressive episodes into results, and longitudinal assessment of cognition alongside evolving depression risk. As a resource, the UK Biobank has general limitations in its representativeness of participants, particularly in terms of general health, ethnicity and socioeconomic status compared to the general population ^24^.

Using a large national cohort, we confirmed remitted depression is a high-risk group for future depression. Age-and sex-matched control participants were at greatest risk of subsequent depression if baseline cognitive performance was poorer, but this relationship was reversed in remitted depression. Ongoing exploration is required to explore methods of targeted intervention based on the specific cognitive profile of remitted depression to support those with previous episodes of depression to stay well.

## Data Availability

Data are available at the UK Biobank (http://www.ukbiobank.ac.uk/).

## CLINICAL IMPLICATIONS

Personalised risk factor assessment for depression is required. Our findings suggest this may be dependent on depression history. Those with no previous history of depression are at higher risk of future depression when cognitive performance is lower at baseline. Remitted depression is a high-risk group for future depression, and those with relatively higher cognitive performance may be more likely to self-report future depressive symptoms.

## ACKNOWLEDGMENTS

We wish to thank Celeste McCracken for her support with data manipulation.

---

**Figure 1**

UK Biobank pipeline as used for this analysis. Figure created in Biorender

**Figure 2**

Future depression expressed as risk </=1. Cognitive scores expressed as a z-score. Confidence intervals around the mean risk shown.

(A) composite score across all dimensions;

(B) memory scores (based on paired associate learning scores)

(B) executive functioning scores (based on digit span, TMT B-A, tower arranging scores);

(C) reasoning scores (based on fluid intelligence, matrix pattern puzzles scores);

(D) processing speed scores (based on reaction time, symbol digit substitution scores).

**Figure 3**

(A) Value = P value where p <0.05 is significant, shown corrected for FDR for main effect of region and for region*group interaction;

(B) Figure created in https://Biorender.com

