## Supplementary Material for "Cognition and Future Depression: Associations with Risk in Those With and Without a History of Depression"

### Supplementary Methods

#### MRI methods

The UK Biobank team generated the brain imaging data which was accessible to researchers.

The exclusion criteria established by the UK Biobank team for MRI scanning. include standard MRI safety/quality criteria, such as exclusions for metal implants, recent surgery or health conditions problematic for MRI scanning, such as claustrophobia. Additionally, raw MRI data that had incorrect dimensions, or were corrupted, missing, or otherwise unusable were not processed and available for inclusion.

The brain imaging data were preprocessed by a fully automated processing pipeline primarily centered around FSL software [1]. The imaging modalities within the UK Biobank comprised T1-weighted, T2-weighted flair, diffusion MRI (dMRI), susceptibility-weighted imaging (SWI), task functional MRI timeseries data (tfMRI), and resting-state functional MRI timeseries data (rsfMRI) [2]. T1-weighted MPRAGE and T2-weighted FLAIR volumes were obtained at 1×1×1mm (208×256×256 field of view [FOV] matrix) and 1.05×1×1mm (192×256×256 FOV matrix) respectively. Linear and non-linear registration to MNI152 "nonlinear 6th generation" standard-space was conducted using FMRIB's Linear Registration Tool (FLIRT) and FMRIB's Nonlinear Image Registration Tool (FNIRT) respectively. Additionally, brain extraction was performed using the Brain Extraction Tool (BET), followed by defacing and segmentation into tissue types using FMRIB's Automated Segmentation Tool (FAST). Grey matter was extracted from FAST and the estimation of the grey matter was performed for a total of 139 regions of interest (ROIs) defined by the Harvard-Oxford cortical and subcortical atlases and the Diedrichsen cerebellar atlas. Subcortical volumes were generated by using FMRIB’s Integrated Registration and Segmentation Tool (FIRST) and estimation was done using the population priors on shape and intensity variation across subjects. Image segmentation was performed to identify white matter hyperintensities (WMH). Additionally, periventricular WMH (pWMH) and deep WMH (dWMH), were defined based on subsets of total WM hyperintensities.

Resting state functional MRI (rsfMRI) were also obtained at 2.4×2.4×2.4mm (88×88×64 FOV matrix). On a preprocessed sample of 4162 participants, grouped average independent component analysis (ICA) was carried out using MELODIC [3, 4]. ICA was performed with dimensionality set to 25 and 100, which resulted in 21 and 55 components, respectively, after discarding the noise components. These 21×21 and 55×55 partial correlation matrices were used as measurements of functional connections. The ICA maps were mapped on each participant’s rsfMRI timeseries data in order to acquire one representative node timeseries per ICA component for each subject. These network ‘nodes’ are a measure of within-network functional connectivity. Subject-specific network-matrices (‘edges’) were also extracted from the node timeseries that provide a measure of functional connectivity between the nodes [5].

Previous UK Biobank studies [15, 21] suggested networks of interest (NOI) for cognition in depression include the following 11 components (nodes) based on “triple network” of default mode network (DMN), central executive network (cEN / FPN) and salience network (SN): (i) DMN = 1, 7, 9, 14, 20; (ii) cEN / FPN = 5, 6, 16, 21 (5, 6 and 21 are part DMN, part cEN); (iii) SN = 3, 13 (13 is part DMN, part SN). 55 edges involving these 11 nodes were taken from [*https://www.fmrib.ox.ac.uk/datasets/ukbiobank/group_means/edge_list_d25.txt*](https://www.fmrib.ox.ac.uk/datasets/ukbiobank/group_means/edge_list_d25.txt)).

MRI methods references

#### Codes used for definition of cohorts and antidepressant use within the UK Biobank

FO+HES codes

| **Code** | **Variable** | **Source** |
| --- | --- | --- |
| 130894 | First report of ICD-10 depressive disorder | First Occurrences |
| 130896 | First report of ICD-10 recurrent depressive disorder | First Occurrences |
| 41270 | HES_summary_diagnosis_ICD-10 | Hospital Episode Statistics |

GP codes

| **Code** | **Variable** |
| --- | --- |
| 1465 | H/O: depression |
| 62T1. | Postnatal depressive disorder |
| E112. | Depression: [single major episode] or [agitated] or [endogenous (including first episode)] |
| E1120 | Single major depressive episode, unspecified |
| E1122 | Single major depressive episode, moderate |
| E1123 | Single major depressive episode, severe, without mention of psychosis |
| E1124 | Single major depressive episode, severe, with psychosis |
| E1125 | Single major depressive episode, in partial or unspecified remission |
| E1126 | Single major depressive episode, in full remission |
| E112z | Single major depressive episode NOS |
| E113. | Recurrent depression: [major episode] or [endogenous] |
| E1130 | Recurrent major depressive episodes, unspecified |
| E1132 | Recurrent major depressive episodes, moderate |
| E1133 | Recurrent major depressive episodes, severe, without mention of psychosis |
| E1134 | Recurrent major depressive episodes, severe, with psychosis |
| E1135 | Recurrent major depressive episodes, in partial or unspecified remission |
| E1136 | Recurrent major depressive episodes, in full remission |
| E1137 | Recurrent depression |
| E113z | Recurrent major depressive episode NOS |
| E11z2 | See X00SU |
| E130. | Reactive depressive psychoses |
| E135. | See X00SQ |
| E2003 | See X00Sb |
| E204. | (Neurotic depression reactive type) or (postnatal depression) |
| .E222 | See X00SR |
| E2B.. | Depressive disorder NEC |
| E2B0. | Postviral depression |
| E2B1. | Chronic depression |
| Eu32. | See XE1Y0 |
| Eu321 | [X]Moderate depressive episode |
| Eu322 | [X] Severe depressive episode without psychotic symptoms: (& [single episode agitated depression] or [single episode major depression] or [single episode vital depression]) |
| Eu323 | [X] Severe depressive episode with psychotic symptoms: (& single episode of [major depression] or [psychogenic depressive psychosis] or [psychotic depression] or [reactive depressive psychosis]) |
| Eu32y | [X] Depression: [other episodes] or [atypical] or [single episode masked NOS] |
| Eu32z | [X] (Depression: [episode, unspecified] or [NOS (& reactive)] or [depressive disorder NOS] |
| Eu325 | See XSGok |
| Eu326 | See XSGol |
| Eu327 | See XSGom |
| Eu328 | See XSGon |
| Eu33. | [X]Recurrent depressive disorder (& [episodes of depressive reaction] or [episodes of psychogenic depression] or [episodes of reactive depression] or [seasonal depressive disorder]) |
| Eu331 | [X]Recurrent depressive disorder, current episode moderate |
| Eu332 | [X]Depression without psychotic symptoms: [recurrent: [major] or [manic-depressive psychosis, depressed type] or [vital] or [current severe episode]] or [endogenous] |
| Eu333 | [X] Depression with psychotic symptoms: [recurrent: (current episode severe) or (manic-depressive, depressed type) or (severe major episodes) or (severe episodes)] or [endogenous] |
| Eu334 | [X]Recurrent depressive disorder, currently in remission |
| Eu33y | [X]Other recurrent depressive disorders |
| Eu33z | [X] Depression recurrent: [unspecified] or [monopolar NOS] |
| .E35 | See XE1aY |
| Eu412 | [X]Mixed anxiety and depressive disorder |
| X00SO | Depressive disorder |
| X00SQ | Agitated depression |
| X00SR | Endogenous depression |
| X00SS | Endogenous depression first episode |
| X00SU | Masked depression |
| X00Sb | Mixed anxiety and depressive disorder |
| X40Dm | Severe postnatal depression |
| XE1Y0 | Single major depressive episode |
| XE1Y1 | Recurrent major depressive episodes |
| XE1YC | Reactive depression |
| XE1aY | Depression: [reactive (neurotic)] or [postnatal] |
| XM1GC | Endogenous depression - recurrent |
| XE1ZY | [X]Severe depressive episode without psychotic symptoms |
| XE1ZZ | [X]Severe depressive episode with psychotic symptoms |
| XE1Za | [X]Other depressive episodes |
| XE1Zb | [X]Depressive episode, unspecified |
| XE1Zc | [X]Recurrent depressive disorder |
| XE1Zd | [X]Recurrent depressive disorder, current episode severe without psychotic symptoms |
| XE1Ze | [X]Recurrent depressive disorder, current episode severe with psychotic symptoms |
| XE1Zf | [X]Recurrent depressive disorder, unspecified |
| XSGok | Major depression |
| XSGol | Moderate major depression |
| XSGom | Severe major depression without psychotic features |
| XSGon | Severe major depression with psychotic features |

Probable depression (UKB derived algorithm) codes

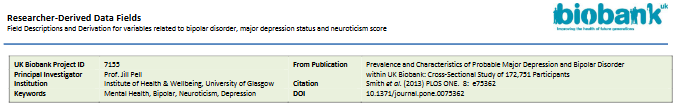

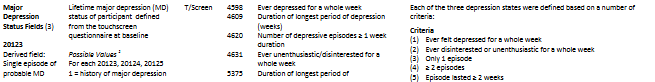

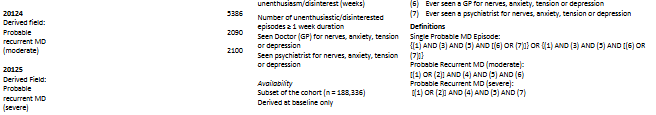

Codes used to ascertain antidepressant use within the UK Biobank (tabular data):

| **Variable name** | **Code** | **Generic name (if different)** | **Class of antidepressant** |
| --- | --- | --- | --- |
| mirtazapine | 1141152732 |  | NASSA |
| zispin 30mg tablet | 1141152736 |  | NASSA |
| mianserin | 1140879556 |  | NASSA |
| zyban 150mg m/r tablet | 1141176858 | Bupropion | Antidepressant |
| tranylcypromine | 1140867914 |  | MAOi |
| parnate 10mg tablet | 1140867916 | Tranylcypromine | MAOi |
| moclobemide | 1140867920 |  | MAOi |
| manerix 150mg tablet | 1140867922 | Moclobemide | MAOi |
| phenelzine | 1140910704 |  | MAOi |
| reboxetine | 1141151978 |  | Noradrenaline reuptake inhibitor |
| edronax 4mg tablet | 1141151982 | Reboxetine | Noradrenaline reuptake inhibitor |
| trazodone | 1140879634 |  | Serotonin modulators |
| nefazodone | 1140917460 |  | Serotonin modulators |
| venlafaxine | 1140916282 |  | SNRI |
| nortriptyline | 1140867818 |  | SNRI |
| clomipramine | 1140879620 |  | SNRI |
| efexor 37.5mg tablet | 1140916288 | Venlafaxine | SNRI |
| citalopram | 1140921600 |  | SSRI |
| fluoxetine | 1140879540 |  | SSRI |
| sertraline | 1140867878 |  | SSRI |
| paroxetine | 1140867888 |  | SSRI |
| escitalopram | 1141180212 |  | SSRI |
| seroxat 20mg tablet | 1140882236 | Paroxetine | SSRI |
| prozac 20mg capsule | 1140867876 | Fluoxetine | SSRI |
| cipralex 5mg tablet | 1141190158 | escitalopram | SSRI |
| lustral 50mg tablet | 1140867884 | Sertraline | SSRI |
| fluvoxamine | 1140879544 |  | SSRI |
| felicium 20mg capsule | 1141170866 | Fluoxetine | SSRI |
| oxactin 20mg capsule | 1141174756 | Fluoxetine | SSRI |
| ranflutin 20mg capsule | 1141185124 | Fluoxetine | SSRI |
| amitriptyline | 1140879616 |  | TCA |
| imipramine | 1140879630 |  | TCA |
| dothiepin | 1140879628 |  | TCA |
| butriptyline | 1140856074 | Butriptyline | TCA |
| evadyne 25mg tablet | 1140856076 | Butriptyline | TCA |
| maprotiline | 1140879552 |  | TCA |
| desipramine | 1140879624 |  | TCA |
| protriptyline | 1140879632 |  | TCA |
| thaden 25mg capsule | 1141171824 | Dosulepin | TCA |

#### Analytic models relevant for each research question as pre-specified

Primary analyses:

Are cognition and depressive relapse associated in remitted depression (RD), and how does this differ from never-depressed (ND) population

- Full sample: Depressive episode after imaging visit ~ Cognitive scores (*total score*)
- ND vs RD: Depressive episode after imaging visit ~ Group*Cognitive scores (*total score*) – PRIMARY OUTCOME

Secondary analyses:

Are there any significant differences between remitted depression and control cohorts for average grey matter volume in key regions of triple network, and do any differences have an impact on (i) concurrent cognitive scores and (ii) relapse

- ND vs RD: Cognitive scores (total score) ~ Group*structural differences
- ND vs RD: Depressive outcome after imaging visit ~ Group*structural differences

Are previous associations found between resting state functional connectivity (rsFC) of Triple Network and cognition in depressed volunteers reproduced in this RD population?

- ND vs RD: Cognitive scores (total score) ~ Group*rsFC (*for networks of interest – amplitudes (nodes) & edges*)

Is rsFC within the Triple Network and depressive relapse associated in RD, and how does this differ from ND population?:

- Full sample: Depressive episode after imaging visit ~ rsFC *(triple network nodes & edges)*
- ND vs RD: Depressive episode after imaging visit ~ Group*rsFC (*triple network nodes & edges*) – PRIMARY OUTCOME

#### Potential confounds (controlled for in regression analyses)

- Demographics (baseline where crystallised to capture all data or imaging visit tabular data)
  - sex (M/F) (31.0 TD) – part of matching process
  - age in years at time of assessment (21003.2 TD) – part of matching process
    - Pre-processed into age^2^ and age^3^, and age × sex, age^2^ × sex for imaging analyses (in addition to matching for age and sex)
  - dates of assessment (53.0/2 TD)
  - assessment centre site (54.2 TD)
  - self-identified ethnic group: White, Black / Black British, Asian & Mixed & Other (21000.0 TD)
- SES / education (baseline where crystallised or imaging visit)
  - self-report of highest qualification (university degree / A levels / GCSEs / CSE / NVQ / professional quals / none) (6138.0 TD (array of 7))
  - neighbourhood deprivation level at recruitment (189)
    - Pre-processed into quintiles; 1 = least deprived and 5 = most deprived)
- Imaging – for imaging analyses *(imaging visit tabular data)*
  - mean rfmri head motion, averaged across space and time points (25741.2.0 TD)
  - structural MRI
    - total brain volume (normalised for head size, 25009.2.0 TD)
    - GM volume (normalised for head size, 25005.2.0 TD)
    - WM volume (normalised for head size, 25007.2.0 TD)
    - CSF volume (normalised for head size, 25003.2.0 TD)
- General health (imaging visit tabular data, FO):
  - BMI (21001.2 TD)
  - smoking (20116.2 TD)
    - Pre-processed into previous and current vs never at imaging
  - before imaging “First Occurence” of common conditions (neurology, cardiovascular, malignancy)
    - G40-G41 epilepsy (131048) (FO)
    - G45-G46 cerebrovascular disease (131056, 131058) (FO)
    - I25 chronic ischaemic heart disease (131306) (FO)
    - Self-reported malignancies (134.2.0 TD)
      - Pre-processed into never vs 1 or more cancers before imaging OR baseline
- Mental health (MHFU, imaging visit tabular data)
  - substance use
    - alcohol consumption frequency (1558.0/2 TD)
      - Pre-processed as units per week

#### Health exclusions (participants excluded if occur at or within 2 years after imaging)

- “First Occurrence” (FO) of dementia, degenerative and demyelinating disorders (G30-G37 Alzheimer’s disease, degenerative disorders, demyelinating disorders (131036; 131038; 131040; 131042; 131044; 131046)(FO)
- G10 Huntington’s disease (131012) (FO)
- G20 Parkinson’s disease (131022) (FO)
- I60 subarachnoid haemorrhage (131360), I61 intracerebral haemorrhage (131362), I63 cerebral infarction (131366) (FO)

#### Prior checks to confirm validity of imputation

1. Load cognitive dataset & transfer across age and sex

z_cog_trial <- readRDS("~/OneDrive - Nexus365/Postdoc_funding/GoB_work/UKB/Inputs_derived/cognition_standardised_scores_fullUKBsample_Feb24.rds")

preproc_demo_slimmed<- readRDS("Inputs_derived/UKB_remitteddep_cog_demographics_preprocessed_Feb24.rds")

#z_cog_trial <- merge(z_cog_trial, preproc_demo_slimmed[c("f.eid", "Sex", "Age_ass_V2")], by = "f.eid", all.x = TRUE)

z_cog_trial <- merge(z_cog_trial, preproc_demo_slimmed, by = "f.eid", all.x = TRUE)

2. Multiple imputation and trial modelling & then check SD across the imputed datasets

\# Load necessary libraries / packages

library(mice)

library(pool)

library(dplyr)

library(nnet)

library(lme4)

\## GROUP A (FO+HES)

\# Step 1: Impute missing data

\# Step 1.1: Remove rows with no data (all empty rows except f.eid/pt id)

z_cog_trial <- z_cog_trial[!apply(is.na(z_cog_trial[-1]), 1, all),]

\# Check the filtered dataset (remove the "stop" line if it looks correct)

#stop("Check the filtered dataset")

\# Step 1.2: Impute missing values

\# Create a function to z-score a column

zscore <- function(x) {

(x - mean(x, na.rm = TRUE)) / sd(x, na.rm = TRUE)

}

\# Columns to be imputed

cols_to_impute <- c(

"reaction_V2_inverse_normal_scores",

"correct_symbol_digit_matches",

"fluid_intell_V2",

"matrix_pattern_puzzles_solved",

"numeric_cog_V2",

"TMTB_MINUS_TMTA_V2_inverse_normal_scores",

"correct_tower_arranging",

"number_correct_paired_associates_V2_inverse_normal_scores"

)

\# Columns to use for imputation

impute_columns <- cols_to_impute

\# Convert the columns to numeric vectors - actually think all cog variables in raw data are raw anyway

#for (col in cols_to_impute) {

#z_cog_trial[[col]] <- as.numeric(z_cog_trial[[col]])

#}

\# Auxiliary variables to improve imputation (but not for imputing)

auxiliary_vars <- c("Sex", "Age_ass_V2")

\# Create an imputation model (first iteration works without auxiliary variables, or can then add them directly into imputation below (hashtagged out if auxiliaries are in model itself as in iteration two)

#imputation_model <- mice(z_cog_trial[impute_columns],

#method = "pmm",

#m = 20, # Number of imputations

#maxit = 50)

\# Convert f.eid columns in both datasets to character

z_cog_trial$f.eid <- as.character(z_cog_trial$f.eid)

preproc_demo_slimmed$f.eid <- as.character(preproc_demo_slimmed$f.eid)

\# Convert data.table to data.frame

preproc_demo_slimmed_df <- as.data.frame(preproc_demo_slimmed)

\# Perform a left join using dplyr

merged_data <- left_join(

select(z_cog_trial, f.eid, all_of(impute_columns)),

select(preproc_demo_slimmed_df, f.eid, Sex, Age_ass_V2),

by = "f.eid"

)

\# Ensure merged_data is a data frame

merged_data <- as.data.frame(merged_data)

\# Specify which variables to impute

impute_vars <- c(

"reaction_V2_inverse_normal_scores",

"correct_symbol_digit_matches",

"fluid_intell_V2",

"matrix_pattern_puzzles_solved",

"numeric_cog_V2",

"TMTB_MINUS_TMTA_V2_inverse_normal_scores",

"correct_tower_arranging",

"number_correct_paired_associates_V2_inverse_normal_scores"

)

\# Create an imputation model

imputation_model <- mice(

data = merged_data,

method = "pmm",

m = 20, # Number of imputations

maxit = 50,

impute = impute_vars,

blockPredictor = list(

c(1, 2, 3, 4, 5, 6, 7, 8), # Predictor block for cognitive variables

c(9, 10) # Predictor block for auxiliary variables (Sex, Age_ass_V2)

)

)

iter imp variable

1	1 correct_symbol_digit_matches fluid_intell_V2 matrix_pattern_puzzles_solved numeric_cog_V2 correct_tower_arranging Sex Age_ass_V2

1	2 correct_symbol_digit_matches fluid_intell_V2 matrix_pattern_puzzles_solved numeric_cog_V2 correct_tower_arranging Sex Age_ass_V2

1	3 correct_symbol_digit_matches fluid_intell_V2 matrix_pattern_puzzles_solved numeric_cog_V2 correct_tower_arranging Sex Age_ass_V2

1	4 correct_symbol_digit_matches fluid_intell_V2 matrix_pattern_puzzles_solved numeric_cog_V2 correct_tower_arranging Sex Age_ass_V2

1	5 correct_symbol_digit_matches fluid_intell_V2 matrix_pattern_puzzles_solved numeric_cog_V2 correct_tower_arranging Sex Age_ass_V2

1	6 correct_symbol_digit_matches fluid_intell_V2 matrix_pattern_puzzles_solved numeric_cog_V2 correct_tower_arranging Sex Age_ass_V2

1	7 correct_symbol_digit_matches fluid_intell_V2 matrix_pattern_puzzles_solved numeric_cog_V2 correct_tower_arranging Sex Age_ass_V2

1	8 correct_symbol_digit_matches fluid_intell_V2 matrix_pattern_puzzles_solved numeric_cog_V2 correct_tower_arranging Sex Age_ass_V2

1	9 correct_symbol_digit_matches fluid_intell_V2 matrix_pattern_puzzles_solved numeric_cog_V2 correct_tower_arranging Sex Age_ass_V2

1	10 correct_symbol_digit_matches fluid_intell_V2 matrix_pattern_puzzles_solved numeric_cog_V2 correct_tower_arranging Sex Age_ass_V2

1	11 correct_symbol_digit_matches fluid_intell_V2 matrix_pattern_puzzles_solved numeric_cog_V2 correct_tower_arranging Sex Age_ass_V2

1	12 correct_symbol_digit_matches fluid_intell_V2 matrix_pattern_puzzles_solved numeric_cog_V2 correct_tower_arranging Sex Age_ass_V2

1	13 correct_symbol_digit_matches fluid_intell_V2 matrix_pattern_puzzles_solved numeric_cog_V2 correct_tower_arranging Sex Age_ass_V2

1	14 correct_symbol_digit_matches fluid_intell_V2 matrix_pattern_puzzles_solved numeric_cog_V2 correct_tower_arranging Sex Age_ass_V2

1	15 correct_symbol_digit_matches fluid_intell_V2 matrix_pattern_puzzles_solved numeric_cog_V2 correct_tower_arranging Sex Age_ass_V2

1	16 correct_symbol_digit_matches fluid_intell_V2 matrix_pattern_puzzles_solved numeric_cog_V2 correct_tower_arranging Sex Age_ass_V2

1	17 correct_symbol_digit_matches fluid_intell_V2 matrix_pattern_puzzles_solved numeric_cog_V2 correct_tower_arranging Sex Age_ass_V2

1	18 correct_symbol_digit_matches fluid_intell_V2 matrix_pattern_puzzles_solved numeric_cog_V2 correct_tower_arranging Sex Age_ass_V2

1	19 correct_symbol_digit_matches fluid_intell_V2 matrix_pattern_puzzles_solved numeric_cog_V2 correct_tower_arranging Sex Age_ass_V2

1	20 correct_symbol_digit_matches fluid_intell_V2 matrix_pattern_puzzles_solved numeric_cog_V2 correct_tower_arranging Sex Age_ass_V2

2	1 correct_symbol_digit_matches fluid_intell_V2 matrix_pattern_puzzles_solved numeric_cog_V2 correct_tower_arranging Sex Age_ass_V2

2	2 correct_symbol_digit_matches fluid_intell_V2 matrix_pattern_puzzles_solved numeric_cog_V2 correct_tower_arranging Sex Age_ass_V2

2	3 correct_symbol_digit_matches fluid_intell_V2 matrix_pattern_puzzles_solved numeric_cog_V2 correct_tower_arranging Sex Age_ass_V2

2	4 correct_symbol_digit_matches fluid_intell_V2 matrix_pattern_puzzles_solved numeric_cog_V2 correct_tower_arranging Sex Age_ass_V2

2	5 correct_symbol_digit_matches fluid_intell_V2 matrix_pattern_puzzles_solved numeric_cog_V2 correct_tower_arranging Sex Age_ass_V2

2	6 correct_symbol_digit_matches fluid_intell_V2 matrix_pattern_puzzles_solved numeric_cog_V2 correct_tower_arranging Sex Age_ass_V2

2	7 correct_symbol_digit_matches fluid_intell_V2 matrix_pattern_puzzles_solved numeric_cog_V2 correct_tower_arranging Sex Age_ass_V2

2	8 correct_symbol_digit_matches fluid_intell_V2 matrix_pattern_puzzles_solved numeric_cog_V2 correct_tower_arranging Sex Age_ass_V2

2	9 correct_symbol_digit_matches fluid_intell_V2 matrix_pattern_puzzles_solved numeric_cog_V2 correct_tower_arranging Sex Age_ass_V2

2	10 correct_symbol_digit_matches fluid_intell_V2 matrix_pattern_puzzles_solved numeric_cog_V2 correct_tower_arranging Sex Age_ass_V2

2	11 correct_symbol_digit_matches fluid_intell_V2 matrix_pattern_puzzles_solved numeric_cog_V2 correct_tower_arranging Sex Age_ass_V2

2	12 correct_symbol_digit_matches fluid_intell_V2 matrix_pattern_puzzles_solved numeric_cog_V2 correct_tower_arranging Sex Age_ass_V2

2	13 correct_symbol_digit_matches fluid_intell_V2 matrix_pattern_puzzles_solved numeric_cog_V2 correct_tower_arranging Sex Age_ass_V2

2	14 correct_symbol_digit_matches fluid_intell_V2 matrix_pattern_puzzles_solved numeric_cog_V2 correct_tower_arranging Sex Age_ass_V2

2	15 correct_symbol_digit_matches fluid_intell_V2 matrix_pattern_puzzles_solved numeric_cog_V2 correct_tower_arranging Sex Age_ass_V2

2	16 correct_symbol_digit_matches fluid_intell_V2 matrix_pattern_puzzles_solved numeric_cog_V2 correct_tower_arranging Sex Age_ass_V2

2	17 correct_symbol_digit_matches fluid_intell_V2 matrix_pattern_puzzles_solved numeric_cog_V2 correct_tower_arranging Sex Age_ass_V2

2	18 correct_symbol_digit_matches fluid_intell_V2 matrix_pattern_puzzles_solved numeric_cog_V2 correct_tower_arranging Sex Age_ass_V2

2	19 correct_symbol_digit_matches fluid_intell_V2 matrix_pattern_puzzles_solved numeric_cog_V2 correct_tower_arranging Sex Age_ass_V2

2	20 correct_symbol_digit_matches fluid_intell_V2 matrix_pattern_puzzles_solved numeric_cog_V2 correct_tower_arranging Sex Age_ass_V2

TRUNCATED FOR SPACE

50	1 correct_symbol_digit_matches fluid_intell_V2 matrix_pattern_puzzles_solved numeric_cog_V2 correct_tower_arranging Sex Age_ass_V2

50	2 correct_symbol_digit_matches fluid_intell_V2 matrix_pattern_puzzles_solved numeric_cog_V2 correct_tower_arranging Sex Age_ass_V2

50	3 correct_symbol_digit_matches fluid_intell_V2 matrix_pattern_puzzles_solved numeric_cog_V2 correct_tower_arranging Sex Age_ass_V2

50	4 correct_symbol_digit_matches fluid_intell_V2 matrix_pattern_puzzles_solved numeric_cog_V2 correct_tower_arranging Sex Age_ass_V2

50	5 correct_symbol_digit_matches fluid_intell_V2 matrix_pattern_puzzles_solved numeric_cog_V2 correct_tower_arranging Sex Age_ass_V2

50	6 correct_symbol_digit_matches fluid_intell_V2 matrix_pattern_puzzles_solved numeric_cog_V2 correct_tower_arranging Sex Age_ass_V2

50	7 correct_symbol_digit_matches fluid_intell_V2 matrix_pattern_puzzles_solved numeric_cog_V2 correct_tower_arranging Sex Age_ass_V2

50	8 correct_symbol_digit_matches fluid_intell_V2 matrix_pattern_puzzles_solved numeric_cog_V2 correct_tower_arranging Sex Age_ass_V2

50	9 correct_symbol_digit_matches fluid_intell_V2 matrix_pattern_puzzles_solved numeric_cog_V2 correct_tower_arranging Sex Age_ass_V2

50	10 correct_symbol_digit_matches fluid_intell_V2 matrix_pattern_puzzles_solved numeric_cog_V2 correct_tower_arranging Sex Age_ass_V2

50	11 correct_symbol_digit_matches fluid_intell_V2 matrix_pattern_puzzles_solved numeric_cog_V2 correct_tower_arranging Sex Age_ass_V2

50	12 correct_symbol_digit_matches fluid_intell_V2 matrix_pattern_puzzles_solved numeric_cog_V2 correct_tower_arranging Sex Age_ass_V2

50	13 correct_symbol_digit_matches fluid_intell_V2 matrix_pattern_puzzles_solved numeric_cog_V2 correct_tower_arranging Sex Age_ass_V2

50	14 correct_symbol_digit_matches fluid_intell_V2 matrix_pattern_puzzles_solved numeric_cog_V2 correct_tower_arranging Sex Age_ass_V2

50	15 correct_symbol_digit_matches fluid_intell_V2 matrix_pattern_puzzles_solved numeric_cog_V2 correct_tower_arranging Sex Age_ass_V2

50	16 correct_symbol_digit_matches fluid_intell_V2 matrix_pattern_puzzles_solved numeric_cog_V2 correct_tower_arranging Sex Age_ass_V2

50	17 correct_symbol_digit_matches fluid_intell_V2 matrix_pattern_puzzles_solved numeric_cog_V2 correct_tower_arranging Sex Age_ass_V2

50	18 correct_symbol_digit_matches fluid_intell_V2 matrix_pattern_puzzles_solved numeric_cog_V2 correct_tower_arranging Sex Age_ass_V2

50	19 correct_symbol_digit_matches fluid_intell_V2 matrix_pattern_puzzles_solved numeric_cog_V2 correct_tower_arranging Sex Age_ass_V2

50	20 correct_symbol_digit_matches fluid_intell_V2 matrix_pattern_puzzles_solved numeric_cog_V2 correct_tower_arranging Sex Age_ass_V2

Warning: Number of logged events: 1

\# Perform imputation

imputed_data <- complete(imputation_model)

\# Create a list to store the individual imputed datasets

imputed_datasets <- list()

\# Access each imputed dataset and store them in the list

for (i in 1:20) {

\# Perform imputation for the current dataset

imputed_data <- complete(imputation_model, action = i)

\# Add the 'f.eid' column to the imputed dataset

#imputed_data$f.eid <- z_cog_trial$f.eid

\# Add auxiliary variables (age and gender) to the imputed dataset

#imputed_data$Sex <- z_cog_trial$Sex

#imputed_data$Age_ass_V2 <- z_cog_trial$Age_ass_V2

\# Add the imputed dataset to the list

imputed_datasets[[i]] <- imputed_data

}

\# Step 1.3: Recalculate the derived z-scores

\# Create a list to store the processed imputed datasets

processed_imputed_datasets <- list()

\# Loop through each imputed dataset

for (i in 1:20) {

imputed_data <- imputed_datasets[[i]]

\# Recalculate the derived z-scores

imputed_data$processing_speed <- rowMeans(

imputed_data[c("reaction_V2_inverse_normal_scores", "correct_symbol_digit_matches")],

na.rm = TRUE

)

imputed_data$processing_speed_zscore <- scale(imputed_data$processing_speed)

imputed_data$reasoning <- rowMeans(

imputed_data[c("fluid_intell_V2", "matrix_pattern_puzzles_solved")],

na.rm = TRUE

)

imputed_data$reasoning_zscore <- scale(imputed_data$reasoning)

imputed_data$executive_function <- rowMeans(

imputed_data[c("numeric_cog_V2", "TMTB_MINUS_TMTA_V2_inverse_normal_scores", "correct_tower_arranging")],

na.rm = TRUE

)

imputed_data$executive_function_zscore <- scale(imputed_data$executive_function)

\# Step 1.4: Set memory_zscore as number_correct_paired_associates_V2_zscore

imputed_data$memory_zscore <- scale(imputed_data$number_correct_paired_associates_V2_inverse_normal_scores)

\# Step 1.5: Calculate the row means of all domains to give a summed score

imputed_data$summed_score <- rowMeans(imputed_data[, c("processing_speed", "reasoning", "executive_function", "number_correct_paired_associates_V2_inverse_normal_scores")])

\# Create z score of summed score

imputed_data$summed_zscores <- scale(imputed_data$summed_score)

\# Store the processed imputed dataset in the list

processed_imputed_datasets[[i]] <- imputed_data

}

saveRDS(processed_imputed_datasets, file = "Inputs_derived/processed_imputed_datasets_Feb24.rds")

\# Extract the lengths of each imputed dataset

dataset_lengths <- sapply(processed_imputed_datasets, function(dataset) nrow(dataset))

\# Print the lengths

print(dataset_lengths)

[1] 502226 502226 502226 502226 502226 502226 502226 502226 502226 502226 502226 502226 502226 502226 502226 502226 502226 502226 502226 502226

\# Check if all lengths are the same

all_lengths_same <- all(dataset_lengths == dataset_lengths[1])

if (all_lengths_same) {

cat("All imputed datasets have the same length.\n")

} else {

cat("Imputed datasets have different lengths.\n")

}

All imputed datasets have the same length.

\# Get list of f.eids from the imputed data (first dataset)

pt_ids_from_first_imputed_dataset <- processed_imputed_datasets[[1]]$f.eid

\# Need this for later 2_match_cohorts

### Supplementary Results

#### Supplementary Figure 1: Flow diagram

UK Biobank participants (n= 501 572)

Participants excluded as no past depression (n=496 783) – pool for controls

Depression in last 15y before imaging (First Occurrences and Hospital Episode Statistics) (n=4789)

Participants excluded as meet criteria for depression at the imaging visit (n=1096)

As above and free from depression at imaging visit (i.e. “remitted”) (n=3693)

Participants missing all cognitive data or functional imaging data (n=802)

As above and with cognitive and functional imaging data (n=2891)

All participants for cognitive analyses including imputation (n=2094)

As above with full set of data for confounding variables (n=2094)

As above and not meeting criteria for a health exclusion (n=2870)

Participants meeting criteria for a health exclusion (n=21)

Participants with complete relevant cognitive data (n=1166)

Matched 1:1 to a control participant from pool (according to age +/- 2y and sex) with complete relevant data and no exclusions

Matched 1:1 to a control participant from pool (according to age +/- 2y and sex) with complete relevant data and no exclusions

Participants missing data for confounding variables (n=776)

Participants with complete relevant MRI data (n=1554)

#### Supplementary Table 1(a): Characteristics at baseline and health factors at imaging of participants with incomplete data for control variables

| **Characteristic** | **Matched control cohort (2870)** | **Primary RD cohort (2870)** |
| --- | --- | --- |
| *Age (years)* | *52.3 (7.33)* | *52.4 (7.20)* |
| *Sex (%): Female; Male* | *1833 (63.9); 1037 (36.1)* | *1833 (63.9); 1037 (36.1)* |
| Townsend Deprivation Index (%) |  |  |
| *1 (= least deprived)* | *1280 (44.6)* | *1195 (41.6)* |
| *2* | *1046 (36.4)* | *1109 (38.6)* |
| *3* | *405 (14.1)* | *409 (14.3)* |
| *4* | *126 (4.39)* | *145 (5.05)* |
| *5 (= most deprived)* | *8 (0.29)* | *10 (0.35)* |
| *Missing* | *5 (0.17)* | *2 (0.07)* |
| Postsecondary Education (%) |  |  |
| *Post university* | 1290 (44.9) | *1252 (43.6)* |
| *A levels* | 383 (13.3) | *384 (13.4)* |
| *O levels* | *586 (20.4)* | *610 (21.3)* |
| *CSEs* | *143 (4.98)* | *147 (5.12)* |
| *NVQ / HND* | *139 (4.84)* | *141 (4.91)* |
| *Other professional* | *127 (4.43)* | *141 (4.91)* |
| *None of the above* | 148 (5.15) | 149 (5.19) |
| *Missing* | 54 (1.88) | 46 (1.60) |
| Ethnicity (%) |  |  |
| *White* | *2769 (96.5)* | *2783 (97.0)* |
| *Black* | *27 (0.94)* | *15 (0.52)* |
| *Other ethnicity groups* | *69 (2.40)* | *61 (2.13)* |
| *Missing* | *5 (0.17)* | *11 (0.40)* |
| Health factors at imaging assessment |  |  |
| *Never smoker* | *1855 (65.3)* | *1690 (59.1)* |
| *Alcohol intake (mean units / week)* | *16.4 (14.2)* | *16.1 (15.3)* |
| *Body mass index (BMI kg / m2) at imaging* | *26.3 (4.44)* | *27.6 (4.97)* |
| Existing conditions at imaging assessment |  |  |
| *Epilepsy* | *26 (0.91)* | *33 (1.15)* |
| *Cerebrovascular disease* | *39 (1.36)* | *49 (1.71)* |
| *Ischaemic heart disease* | *89 (3.10)* | *171 (5.96)* |
| *Previous cancer diagnosis* | *332 (11.6)* | *430 (15.0)* |
| Depression factors |  |  |
| *Close family history of depression at baseline* | *219 (7.6)* | *442 (15.4)* |
| *Antidepressant use at imaging (%)* | *0 (0)* | *981 (34.2)* |

#### Supplementary Table 1(b): Characteristics at baseline and health factors at imaging of participants with complete MRI data

| **Characteristic** | **Matched control cohort (1554)** | **Primary RD cohort (1554)** |
| --- | --- | --- |
| *Age (years)* | 52.6 (7.34) | 52.9 (7.19) |
| *Sex (%): Female; Male* | 923 (59.4); 631 (40.6) | 923 (59.4); 631 (40.6) |
| Townsend Deprivation Index (%) |  |  |
| *1 (= least deprived)* | 722 (46.5) | 672 (43.2) |
| *2* | 566 (36.4) | 602 (38.7) |
| *3* | 209 (13.4) | 207 (13.3) |
| *4* | 54 (3.47) | 67 (4.31) |
| *5 (= most deprived)* | 3 (0.19) | 6 (0.39) |
| Postsecondary Education (%) |  |  |
| *Post university* | 765 (49.2) | 733 (47.2) |
| *A levels* | 206 (13.3) | 213 (13.4) |
| *O levels* | 312 (20.1) | 317 (20.4) |
| *CSEs* | 80 (5.15) | 78 (5.02) |
| *NVQ / HND* | 73 (4.70) | 72 (4.63) |
| *Other professional* | 58 (3.73) | 71 (4.57) |
| *None of the above* | 60 (3.86) | 70 (4.50) |
| Ethnicity (%) |  |  |
| *White* | 1520 (97.8) | 1532 (98.6) |
| *Black* | 8 (0.51) | 3 (0.19) |
| *Other ethnicity groups* | 26 (1.67) | 19 (1.22) |
| Health factors at imaging assessment |  |  |
| *Never smoker* | 999 (64.2) | 877 (56.4) |
| *Alcohol intake (mean units / week)* | 16.5 (14.0) | 16.2 (15.1) |
| *Body mass index (BMI kg / m2) at imaging* | 26.1 (4.28) | 27.2 (4.68) |
| Existing conditions at imaging assessment |  |  |
| *Epilepsy* | 6 (0.39) | 15 (0.97) |
| *Cerebrovascular disease* | 23 (1.48) | 36 (2.31) |
| *Ischaemic heart disease* | 60 (3.86) | 109 (7.01) |
| *Previous cancer diagnosis* | 170 (10.9) | 231 (14.9) |
| Depression factors |  |  |
| *Close family history of depression at baseline* | 219 (7.6) | 442 (15.4) |
| *Antidepressant use at imaging (%)* | 0 (0) | 537 (34.6) |

#### Supplementary Table 2(a): Comparison between remitted depressed cohort with complete data for control variables (n=2094) and matched control cohort (n=2094) for FO+HES cohort

| **Characteristic / health factor** | **Estimate** | **P value** |
| --- | --- | --- |
| *Ethnicity* | 4.93 (chi-sq) | 0.08 |
| *Education* | 5.22 (chi-sq) | 0.51 |
| *Deprivation (Townsend Index)* | 13.9 (chi-sq) | **0.008** |
| *BMI* | 80.8 (F) | **<0.001** |
| *Smoking* | 23.3 (chi-sq) | **<0.001** |
| *Epilepsy* | 0.44 (chi-sq) | 0.51 |
| *Cerebrovascular disease* | 0.93 (chi-sq) | 0.33 |
| *Ischaemic heart disease* | 30.4 (chi-sq) | **<0.001** |
| *Previous cancer* | 5.16 (chi-sq) | **0.02** |
| *Alcohol* | 0.83 (F) | 0.36 |

#### Supplementary Table 2(b): Comparison between remitted depressed cohort with complete data for control variables (n=1123) and matched control cohort (n=1123) for GP cohort

| **Characteristic / health factor** | **Estimate** | **P value** |
| --- | --- | --- |
| *Ethnicity* | 6.58 (chi-sq) | **0.04** |
| *Education* | 8.41 (chi-sq) | 0.21 |
| *Deprivation (Townsend Index)* | 4.61 (chi-sq) | 0.33 |
| *BMI* | 18.5 (F) | **<0.001** |
| *Smoking* | 7.52 (chi-sq) | **<0.001** |
| *Epilepsy* | 0.95 (chi-sq) | 0.33 |
| *Cerebrovascular disease* | 9.41 (chi-sq) | **0.002** |
| *Ischaemic heart disease* | 9.87 (chi-sq) | **0.002** |
| *Previous cancer* | 0.00 (chi-sq) | 1 |
| *Alcohol* | 0.85 (F) | 0.77 |

#### Supplementary Table 2(c): Comparison between remitted depressed cohort with complete data for control variables (n=2534) and matched control cohort (n=2534) for Probable Depression cohort

| **Characteristic / health factor** | **Estimate** | **P value** |
| --- | --- | --- |
| *Ethnicity* | 2.77 (chi-sq) | 0.25 |
| *Education* | 15.4 (chi-sq) | **0.018** |
| *Deprivation (Townsend Index)* | 9.97 (chi-sq) | **0.041** |
| *BMI* | 32.7 (F) | **<0.001** |
| *Smoking* | 14.6 (chi-sq) | **<0.001** |
| *Epilepsy* | 0.12 (chi-sq) | 0.73 |
| *Cerebrovascular disease* | 3.09 (chi-sq) | 0.08 |
| *Ischaemic heart disease* | 1.72 (chi-sq) | 0.19 |
| *Previous cancer* | 2.21 (chi-sq) | 0.14 |
| *Alcohol* | 0.26 (F) | 0.61 |

#### Supplementary Table 3: Behavioural cognitive functioning at imaging (without imputation)

*Summed scores for each cognitive domain and the overall composite measure combining all domains (scores below shown are z scores).*

*Tasks included as follows per domain: processing speed (reaction time, symbol digit substitution), reasoning (fluid intelligence, matrix pattern puzzles), executive function (digit span, TMT B-A, tower arranging), memory (paired associate learning).*

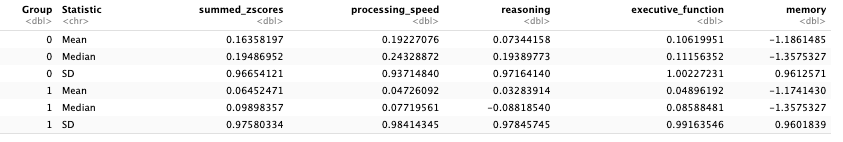

Group 0 = Controls; Group 1 = Remitted depressed cohort (FO+HES)

#### Supplementary Table 4: Differences between RD and matched control primary cohorts when compared with other secondary RD and matched control cohorts

|  | ***Estimate (p value)*** | | | ***Direction if significant (and if similar across cohorts)*** |
| --- | --- | --- | --- | --- |
| ***Baseline measure*** | *FO+HES cohort (n=2094)* | *GP cohort (n=1123)* | *Prob Dep cohort (n=2534)* |  |
| ***Ethnicity (x-sq)*** | 4.93 (p=0.08) | 6.58 (p=0.04) | 2.77 (p=0.25) | More likely to be white in RD cohort than controls (GP significant) |
| ***Townsend (x-sq)*** | 13.9 (p=0.008) | 4.61 (p=0.33) | 13.2 (p=0.01) | Least deprived (TDI 1) higher proportion in controls (FO+HES and Prob Dep significant) |
| ***Education (x-sq)*** | 5.22 (p=0.52) | 8.42 (p=0.21) | 15.4 (p=0.02) | Higher proportion of higher education groups in depressed group (Prob Dep significant) |
| ***White matter (F)*** | 4.00 (p=0.05) | 4.57 (p=0.03) | 1.63 (p=0.20) | WM volume tending to be reduced in RD cohorts (FO+HES borderline and Prob Dep significant) |
| ***Grey matter (F)*** | 6.28 (p=0.012) | 3.63 (p=0.06) | 0.02 (p=0.88) | GM volume tending to be reduced in RD cohorts (FO+HES significant and GP borderline) |
| ***Antidepressants (% in RD cohort)*** | 34.2 | 26.4 | 13.5 | Lower proportion on antidepressants in Prob Dep cohort |

#### Supplementary Table 5(a): Differences with other cohorts in terms of cognitive functioning at the imaging visit

(i) GP cohort versus matched controls

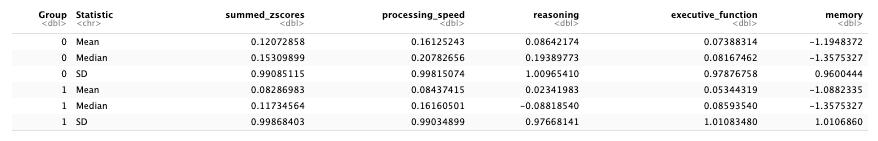

(ii) Probable Depression versus matched controls

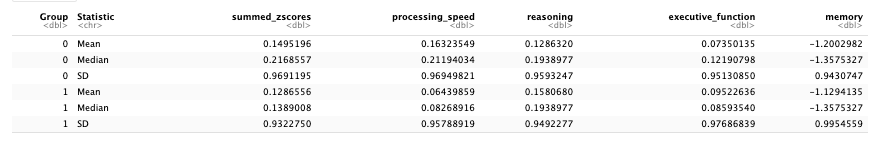

#### Supplementary Table 5(b): Sources of longitudinal data for future depressive episodes (n; %)

For participants with no previous episode of depression (n=179):

| Hospital Episode Statistics | Mental Health Follow Up (record of age at last depression later than age at imaging) | Mental Health Follow Up (current PHQ-9>/=10) | Online Follow Up (self-report of recent depression) |
| --- | --- | --- | --- |
| 19 (10.6%) | 110 (61.4%) | 116 (64.8%) | 173 (97%) |

For participants with remitted depression (n=620):

| Hospital Episode Statistics | Mental Health Follow Up (record of age at last depression later than age at imaging) | Mental Health Follow Up (current PHQ-9>/=10) | Online Follow Up (self-report of recent depression) |
| --- | --- | --- | --- |
| 348 (56.1%) | 442 (71.3%) | 416 (67.1%) | 595 (96.0%) |

*NB. Some participants have > one simultaneous report of depression in follow-up period*

#### Supplementary Table 6: Risk of future depressive episodes and impact of composite cognitive scores at imaging visit

| **Analyses** |  | **Beta (estimate)** | **Standard error** | **Z score** | **P value** |
| --- | --- | --- | --- | --- | --- |
| ***Main model*** | *Effect of composite cognitive score* | -0.263 | 0.111 | -2.374 | 0.018* |
|  | *Composite cognitive score*group* | 0.415 | 0.128 | 3.246 | 0.001* |
| ***Model without group*** | *Effect of composite cognition score* | 0.006 | 0.058 | 0.102 | 0.919 |
| ***Main model with imputation*** | *Effect of composite cognitive score* | -0.167 | 0.078 | -2.144 | 0.032* |
|  | *Composite cognitive score*group* | 0.251 | 0.089 | 2.80 | 0.005* |

#### Supplementary Figure 2: Future depression risk relating to cognition scores at imaging visit for all comers (model without group)

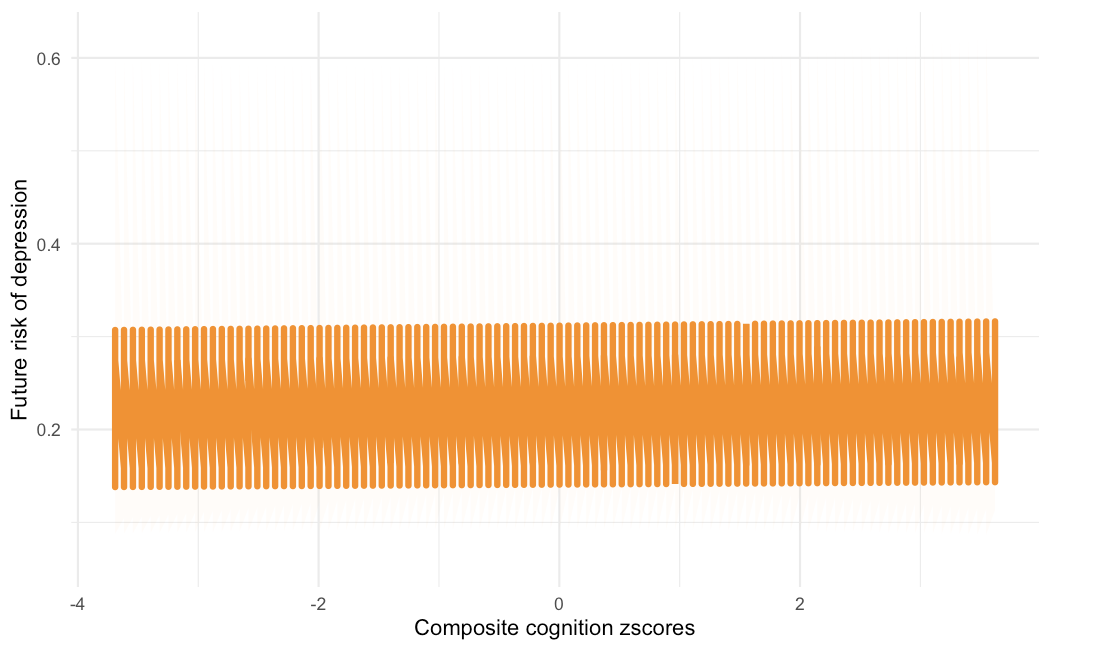

*Future depression expressed as risk </=1. Cognitive scores expressed as a z score. Confidence intervals around the mean risk shown. (A) composite score across all dimensions; (B) memory scores (based on paired associate learning scores) (B) executive functioning scores (based on digit span, TMT B-A, tower arranging scores); (C) reasoning scores (based on fluid intelligence, matrix pattern puzzles scores); (D) processing speed scores (based on reaction time, symbol digit substitution scores)*

#### Supplementary Table 7(a): Risk of future depressive episodes and impact of memory score (n=4188; RD:controls=2094:2094)

| **Analyses** |  | **Beta (estimate)** | **Standard error** | **Z score** | **P value** |
| --- | --- | --- | --- | --- | --- |
| ***Main model*** | *Effect of memory score* | -0.061 | 0.085 | -0.718 | 0.472 |
|  | *Memory score*group* | -0.025 | 0.094 | -0.265 | 0.791 |
| ***Model without group*** | *Effect of memory scores* | -0.081 | 0.050 | -1.624 | 0.104 |

#### Supplementary Table 7(b): Risk of future depressive episodes and impact of executive function (EF) score (n=2395; RD:controls=1174:1221)

| **Analyses** |  | **Beta (estimate)** | **Standard error** | **Z score** | **P value** |
| --- | --- | --- | --- | --- | --- |
| ***Main model*** | *Effect of EF score* | -0.269 | 0.105 | -2.560 | 0.010* |
|  | *EF score*group* | 0.421 | 0.123 | 3.412 | 0.0006* |
| ***Model without group*** | *Effect of EF score* | 0.009 | 0.055 | 0.171 | 0.864 |

#### Supplementary Table 7(c): Risk of future depressive episodes and impact of reasoning score (n=3098; RD:controls=1529:1569)

| **Analyses** |  | **Beta (estimate)** | **Standard error** | **Z score** | **P value** |
| --- | --- | --- | --- | --- | --- |
| ***Main model*** | *Effect of reasoning score* | -0.110 | 0.100 | -1.102 | 0.270 |
|  | *Reasoning score*group* | 0.240 | 0.114 | 2.102 | 0.036* |
| ***Model without group*** | *Effect of reasoning score* | 0.058 | 0.053 | 1.085 | 0.278 |

#### Supplementary Table 7(d): Risk of future depressive episodes and impact of processing speed (PS) score (n=3122; RD:controls=1541:1581)

| **Analyses** |  | **Beta (estimate)** | **Standard error** | **Z score** | **P value** |
| --- | --- | --- | --- | --- | --- |
| ***Main model*** | *Effect of PS score* | -0.185 | 0.101 | -1.832 | 0.067 |
|  | *PS score*group* | 0.253 | 0.116 | 2.182 | 0.029* |
| ***Model without group*** | *Effect of PS score* | -0.037 | 0.053 | -0.729 | 0.466 |

#### Supplementary Table 8: Risk of future depressive episodes and impact of individual cognitive tasks

| **Analyses (all main model)** |  | **Beta (estimate)** | **Standard error** | **Z score** | **P value** |
| --- | --- | --- | --- | --- | --- |
| ***TMTB-A (time; inverse)*** | *Effect of task measure* | -1.966 | 0.077 | -0.255 | 0.798 |
| ***N=*** | *Task measure*group* | -0.104 | 0.087 | 1.194 | 0.232 |
| ***Reaction time (time; inverse)*** | *Effect of task measure* | -0.215 | 0.112 | -1.915 | 0.055 |
|  | *Task measure*group* | 0.168 | 1.342 | 1.252 | 0.211 |
| ***Digit symbol matches (number correct)*** | *Effect of task measure* | -0.180 | 0.101 | -1.79 | 0.074 |
|  | *Task measure*group* | 0.246 | 0.116 | 2.124 | 0.034* |
| ***Fluid intelligence (score)*** | *Effect of task measure* | -0.088 | 0.086 | -1.025 | 0.306 |
|  | *Task measure*group* | 0.216 | 0.099 | 2.190 | 0.029* |
| ***Matrix pattern puzzles (number solved correctly)*** | *Effect of task measure* | -0.149 | 0.096 | -1.537 | 0.124 |
|  | *Task measure*group* | 0.207 | 0.112 | 1.854 | 0.063 |
| ***Numerical cognition (score)*** | *Effect of task measure* | -0.046 | 0.103 | -0.452 | 0.652 |
|  | *Task measure*group* | 0.137 | 0.121 | 1.132 | 0.258 |
| ***Tower arranging (number correct)*** | *Effect of task measure* | -0.260 | 0.105 | -2.489 | 0.013 |
|  | *Task measure*group* | 0.379 | 1.224 | 3.097 | 0.002* |
| ***Paired associates (number correct)*** | *Effect of task measure* | -0.061 | 0.085 | -0.718 | 0.473 |
|  | *Task measure*group* | -0.025 | 0.094 | -0.265 | 0.791 |

#### Supplementary Figure 3: Future depression risk relating to cognition scores at imaging visit for individual tasks

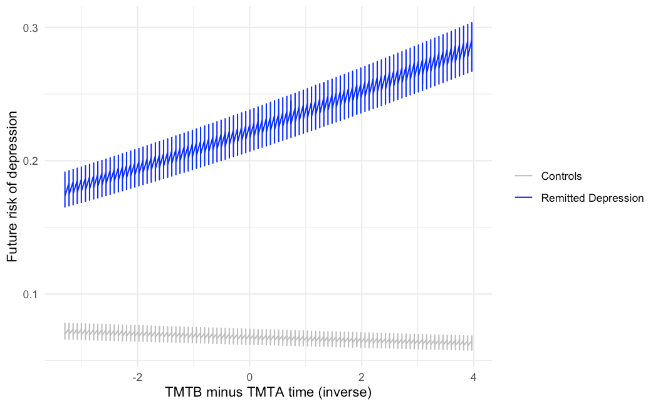

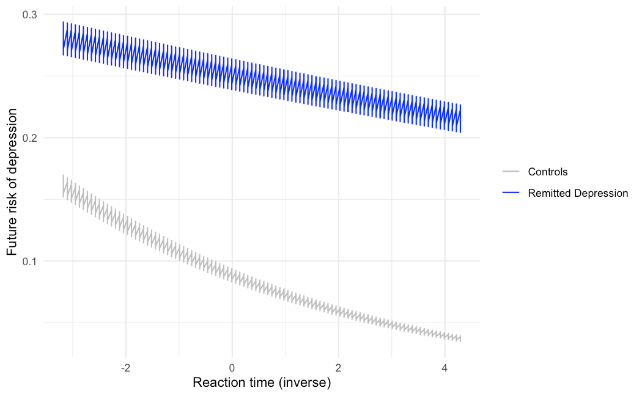

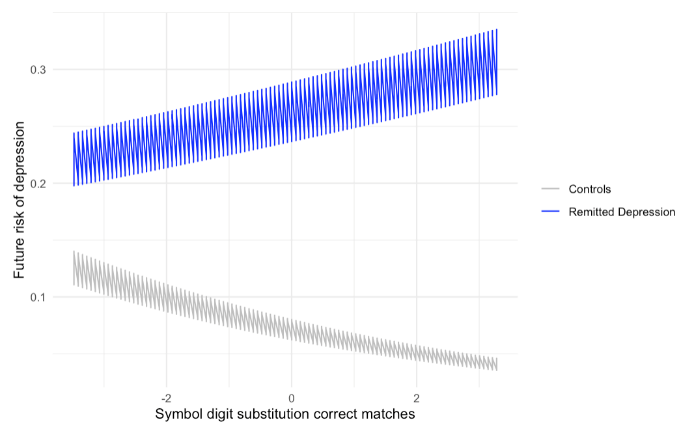

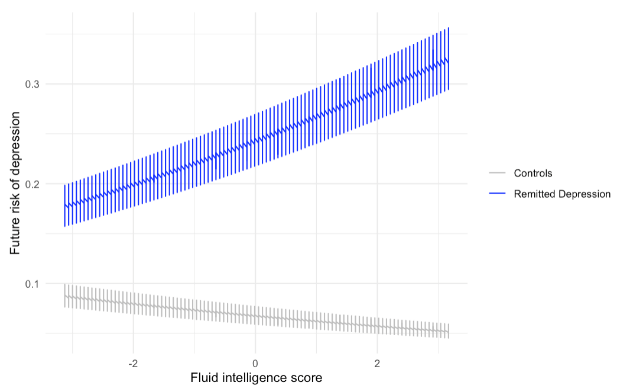

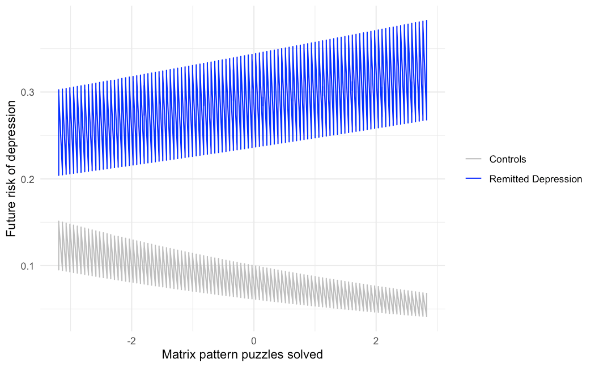

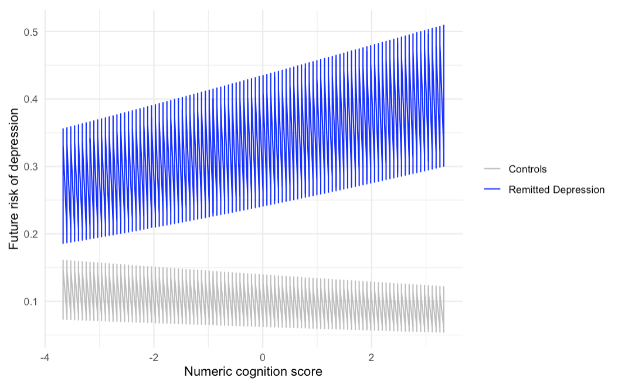

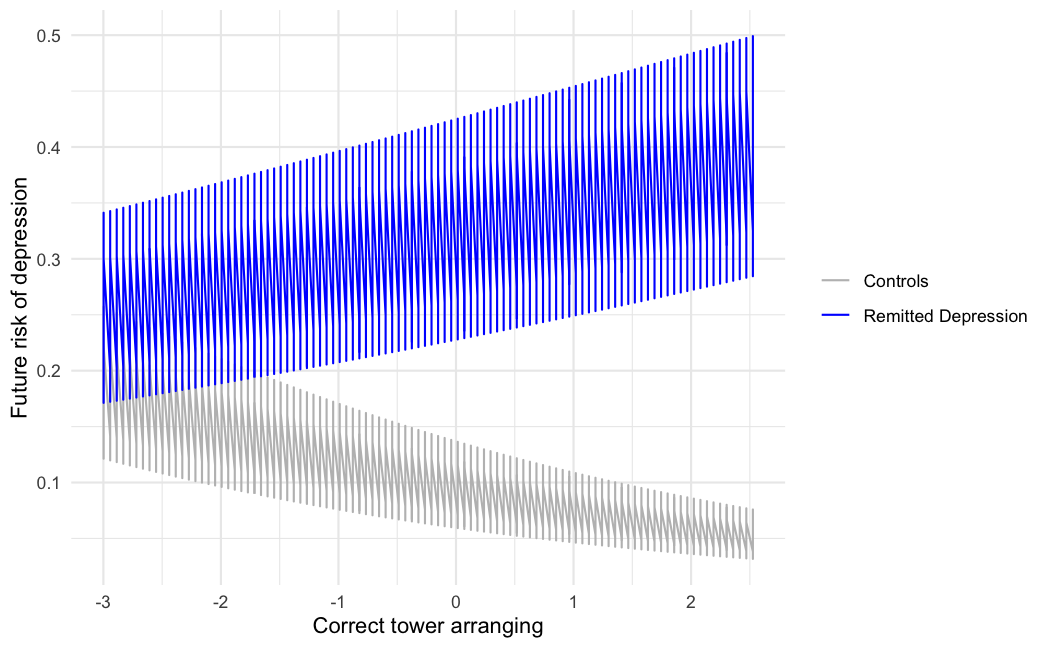

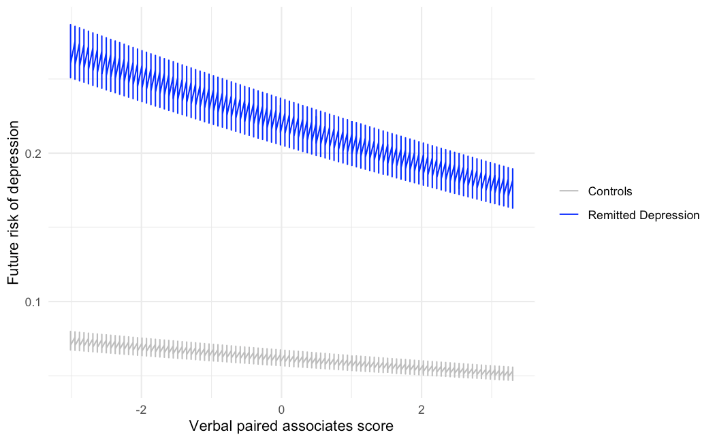

*Future depression expressed as risk </=1. Cognitive scores expressed as a z score. Confidence intervals around the mean risk shown.*

#### Results from Sensitivity Analyses for Primary outcome: characteristics for GP cohort and Probable Depression cohort

Primary RD (using First Occurrences and Hospital Episode Statistics (FO+HES)) and secondary RD cohorts (using GP data; and the “Probable Depression” UKB-derived algorithm data) were similar in terms of demographic, health and imaging parameters. Differences are included in Supplementary Table 2(b) and 2(c) and summarised in Supplementary Table 4. In brief, significant differences comparing RD and control participants using less stringent depressive thresholds were that (i) in the GP cohort, RD participants were more likely to be of white ethnicity than controls; (ii) in the Probable Depression cohort, RD participants were more likely to be more living in deprived postcodes and to be in the higher education group compared to controls. The Probable Depression cohort also had fewer RD participants who were in receipt of antidepressants at the imaging visit (13.5% versus 26.4% (GP) and 34.2% (FO+HES)). However, for both the GP and Probable Depression RD cohort, compared to their control participants, there were no significant differences in performance on cognitive behavioural tasks (baseline summed z scores or on the individual domain level; ps>0.05) (see Supplementary Table 5). This contrasts to that seen in the primary FO+HES RD group, where compared to controls, RD participants had a lower overall performance on cognitive tasks compared to controls.

#### Results from Sensitivity Analyses for Primary outcome: (i) Summed cognitive score impact on future risk of depression for GP cohort:

N=1123 in RD cohort + 1123 in matched controls = 2246.

328 (29%) in RD group and 81 (7%) in control group developed depression longitudinally after imaging visit

Incomplete cognitive data for 959 across both groups (473 in controls and 486 in RD cohort)

The composite cognition measure did not significantly explain group-level differences in the risk of future depression (z=-1.12, p=0.26), and there was no significant interaction between composite cognition score and group (z=0.84, p=0.40). Models without the inclusion of group identified that there was no general association between composite cognition score and future depression across the two groups (p=0.43).

#### Results from Sensitivity Analyses for Primary outcome: (ii) Summed cognitive score results for Probable Depression cohort:

N=2534 in RD cohort + 2534 in matched controls = 5068.

430 (16%) in RD group and 204 in control group developed depression longitudinally after imaging visit

Incomplete cognitive data for 2216 across both groups (1093 in controls and 1123 in RD cohort)

The composite cognition measure did not significantly explain group-level differences in the risk of future depression (z=-1.12, p=0.26), and there was no significant interaction between composite cognition score and group (z=1.10, p=0.27). Models without the inclusion of group identified that there was no general association between composite cognition score and future depression across the two groups (p=0.62).

#### Supplementary Figure 3: Future depression risk relating to composite cognition scores at imaging: (A) using GP records to define past depression; (B) using the Probable Depression algorithm to define past depression

A

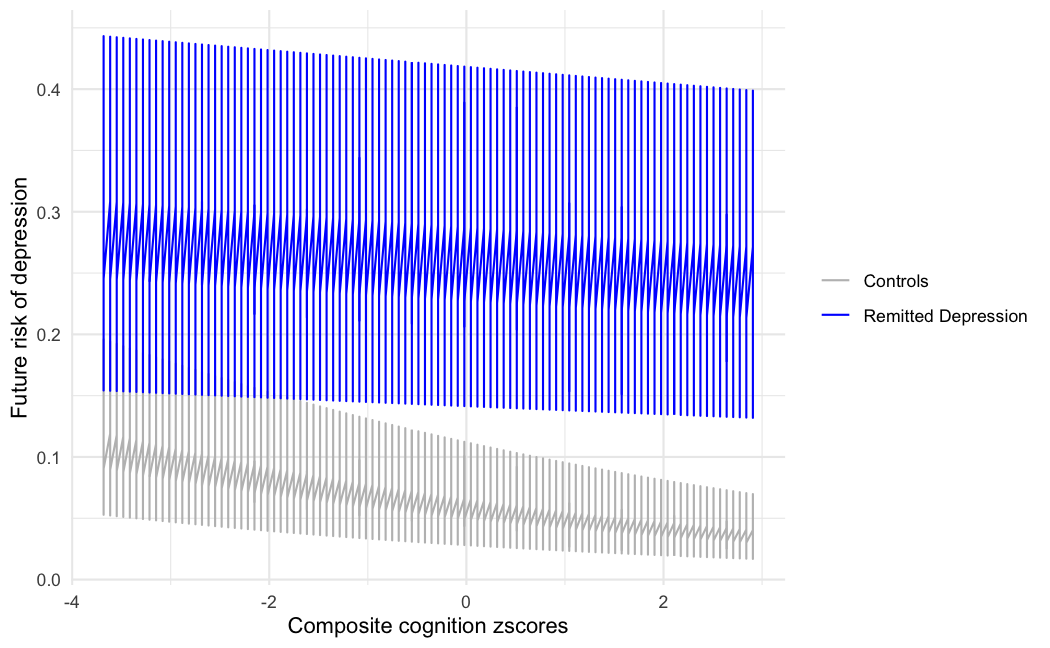

B

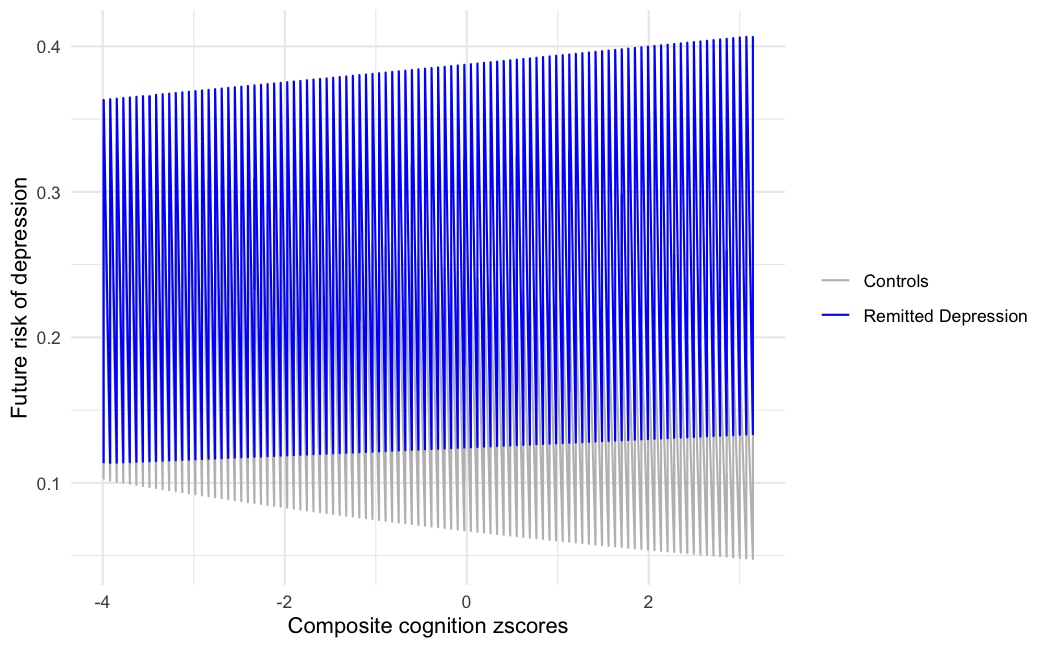

*Future depression expressed as risk </=1. Cognitive scores expressed as a z score. Confidence intervals around the mean risk shown. (A) composite score across all dimensions; (B) memory scores (based on paired associate learning scores) (B) executive functioning scores (based on digit span, TMT B-A, tower arranging scores); (C) reasoning scores (based on fluid intelligence, matrix pattern puzzles scores); (D) processing speed scores (based on reaction time, symbol digit substitution scores)*

#### Supplementary Figure 4: Dimensional comparison across the primary and secondary cohorts

(A) Memory
(i) FO+HES (N=4188)

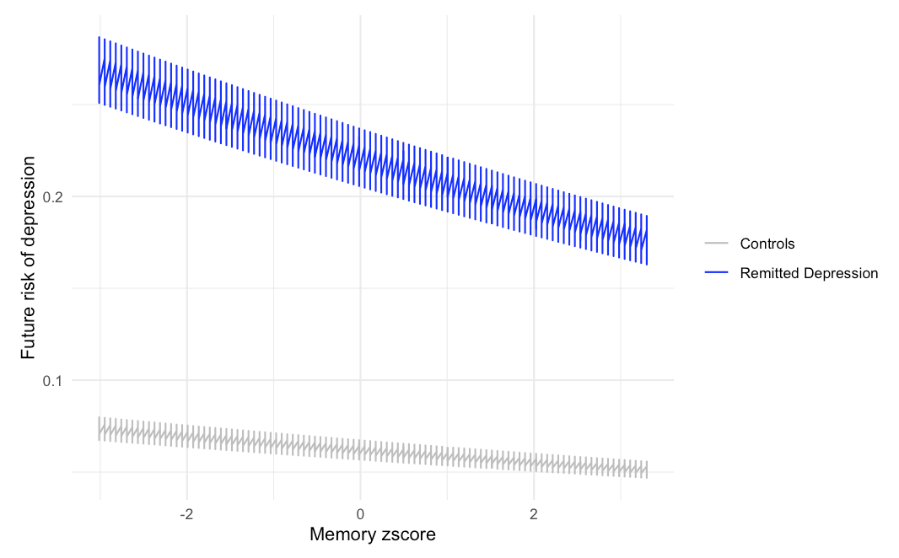

(ii) GP (N=2246)

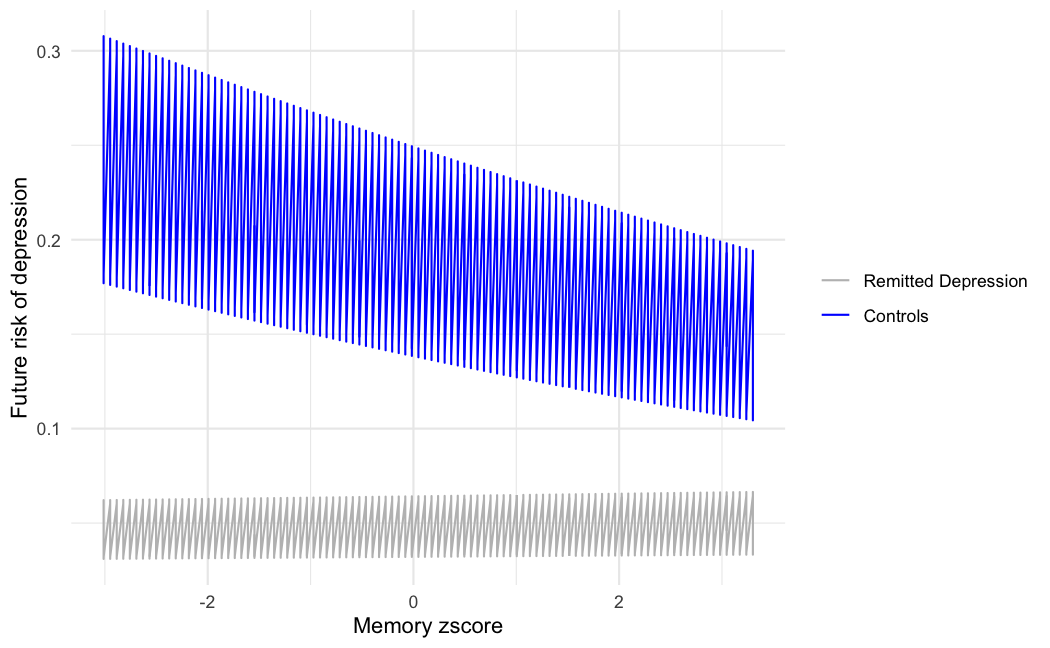

(iii) Probable Depression (N=5068)

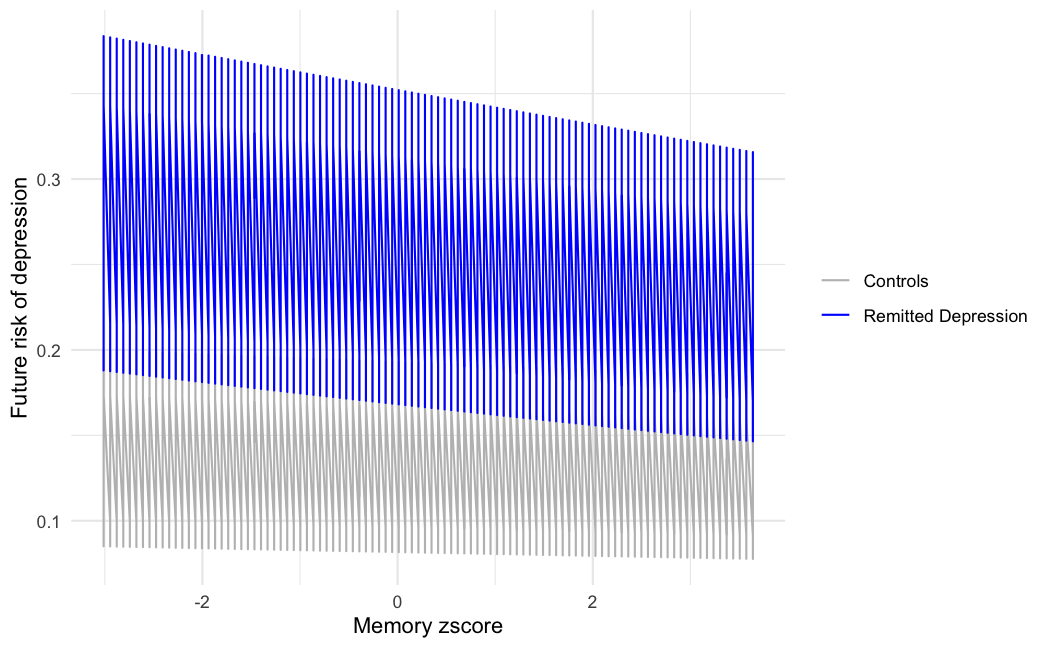

(B) Executive function
(i) FO+HES (N=2395)

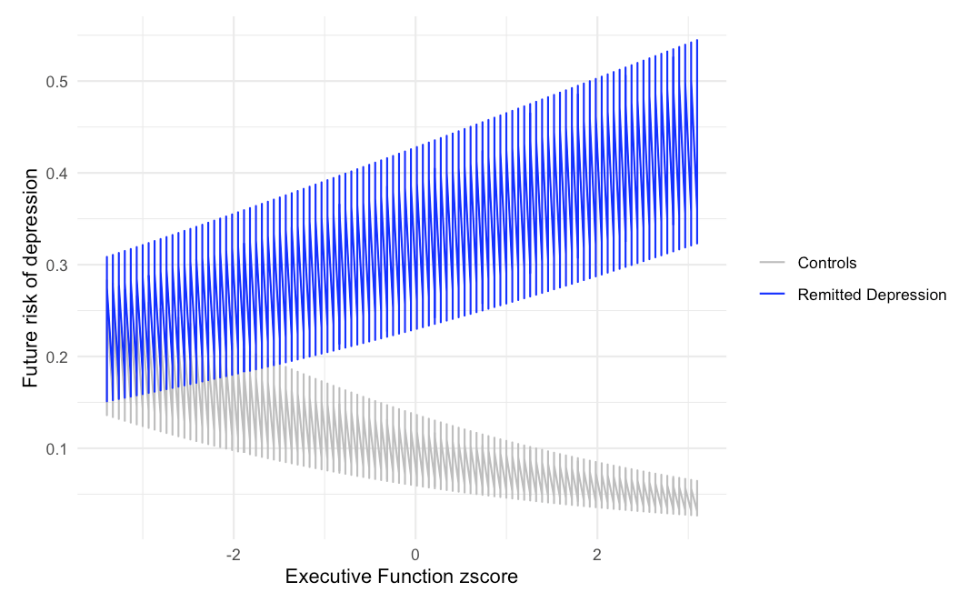

(ii) GP (N=1294)

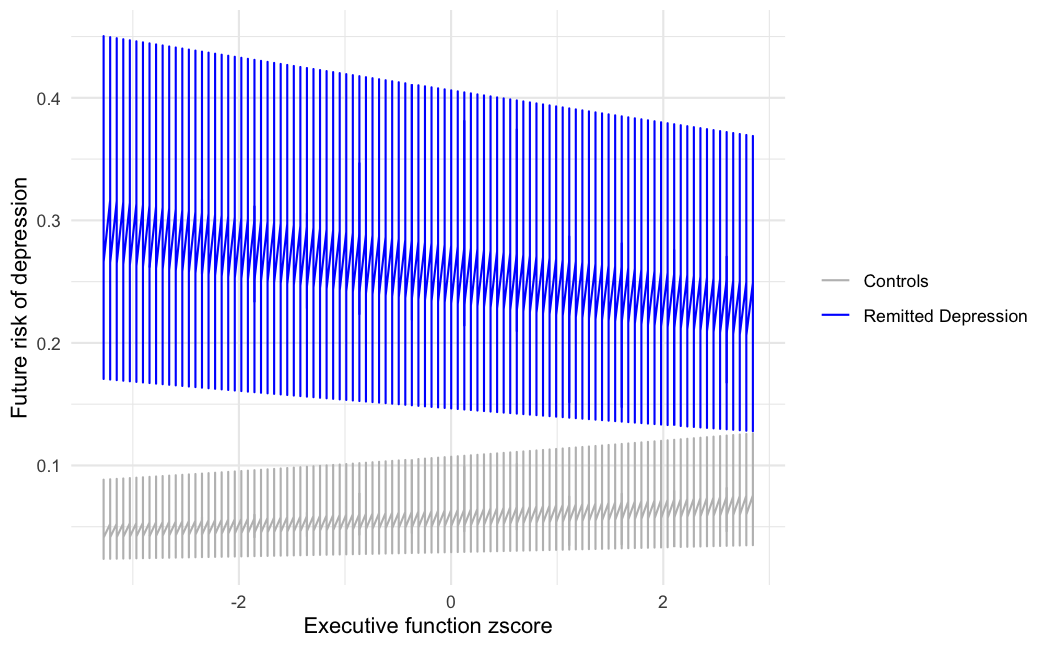

(iii) Probable Depression (N=2886)

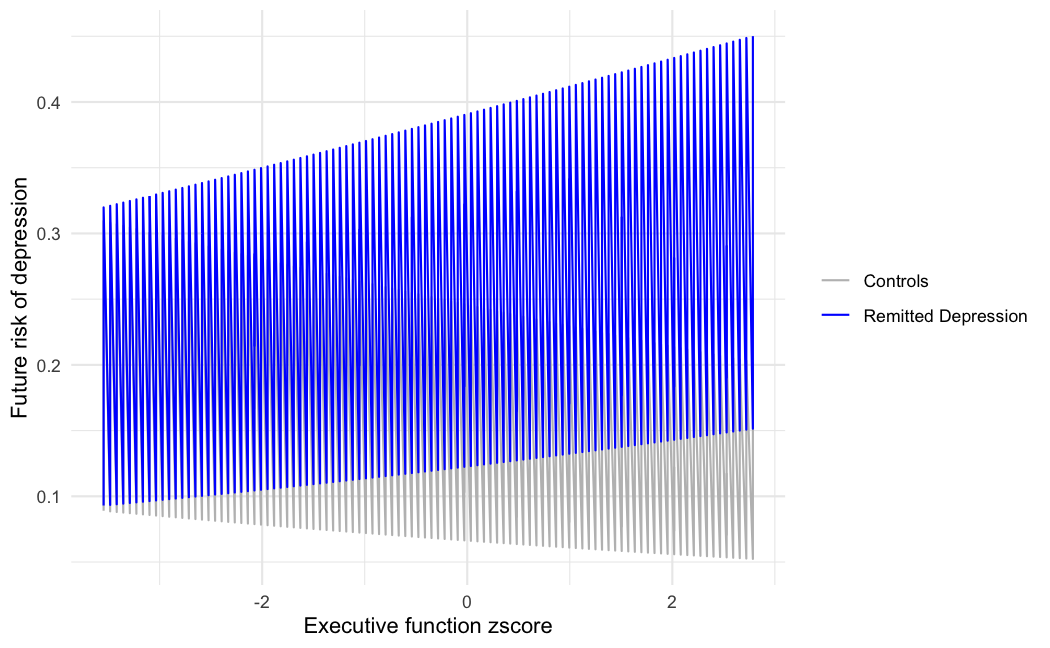

(C) Reasoning
(i) FO+HES (N=3098)

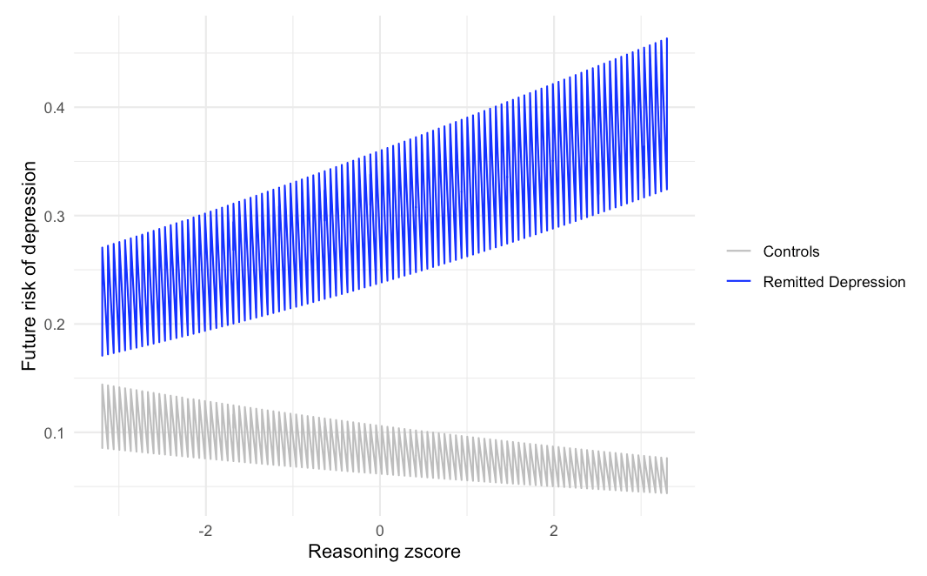

(ii) GP (N=1634)

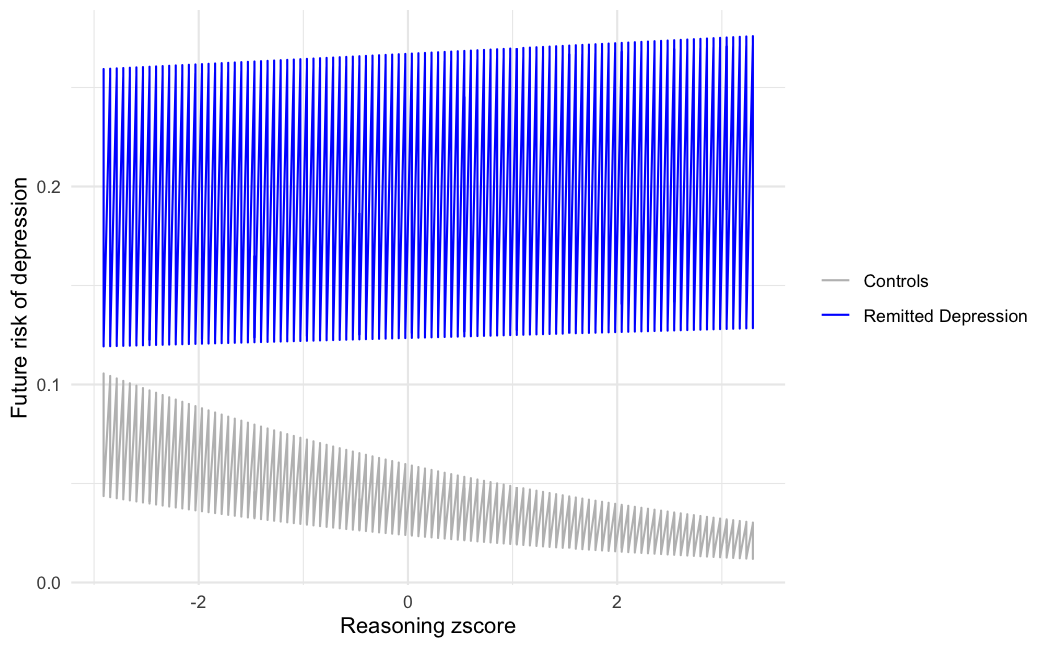

(iii) Probable Depression (N=3777)

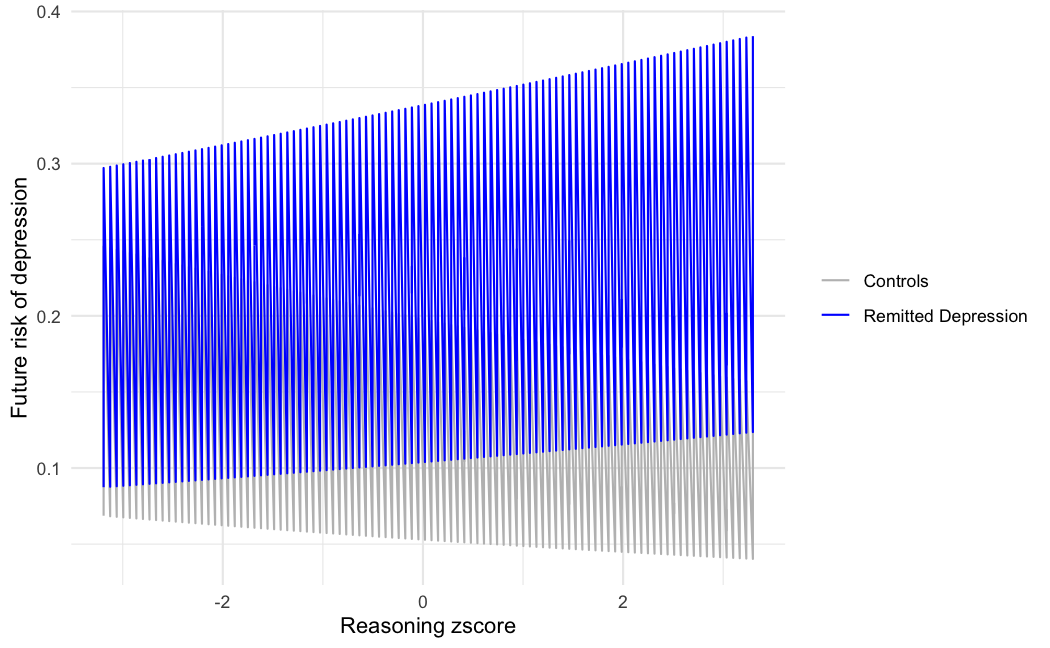

(D) Processing speed
(i) FO+HES (N=3112)

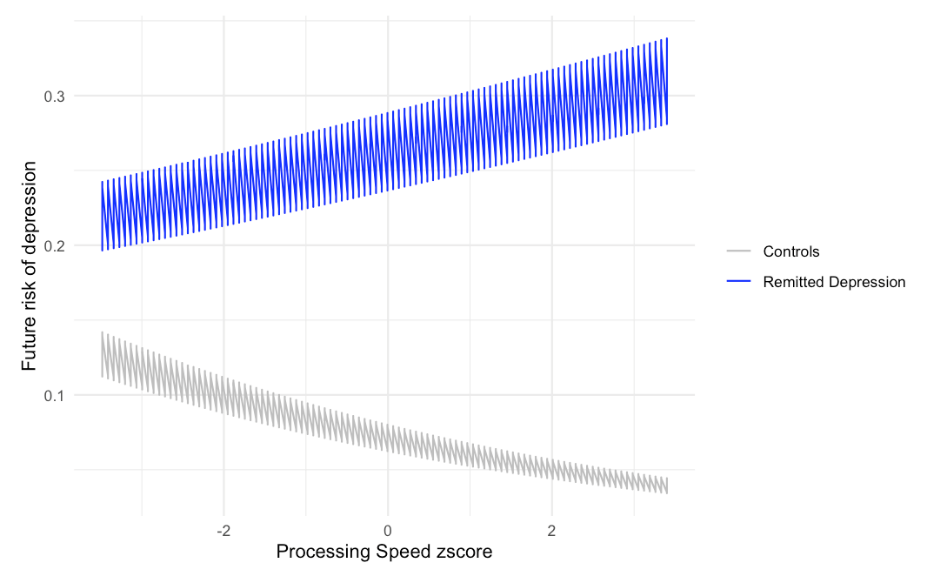

(ii) GP	(N=1647)

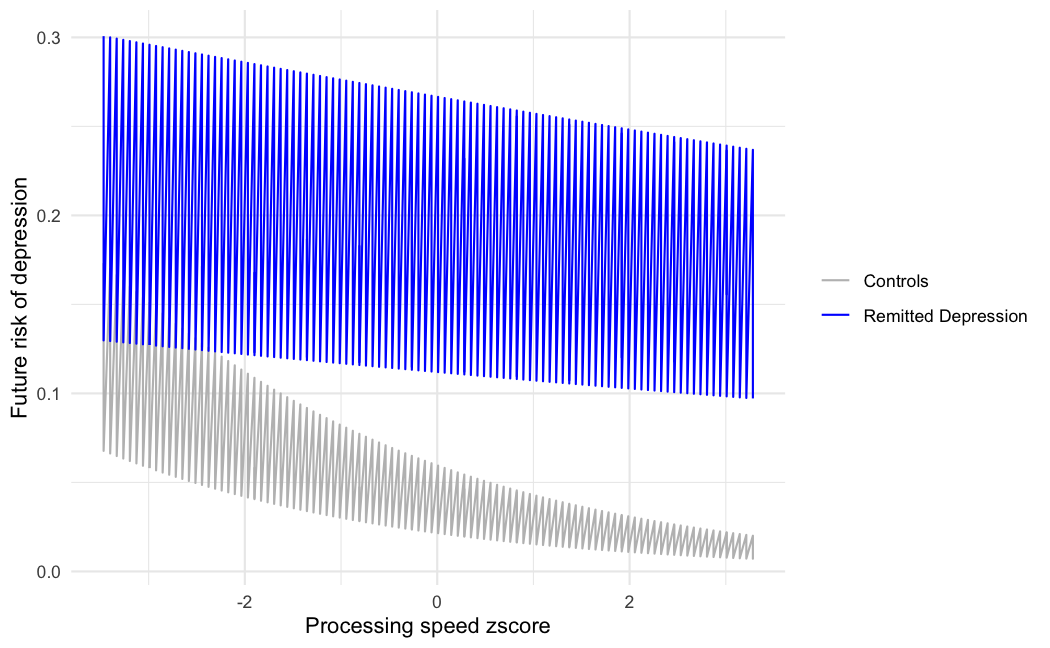

(iii) Probable Depression (N=3792)

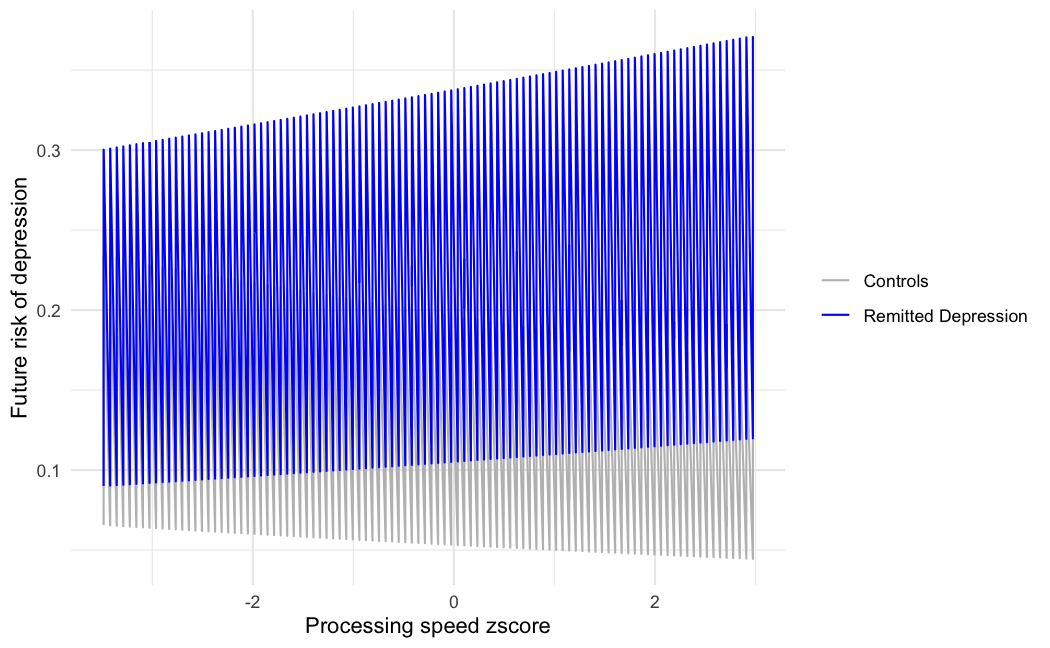

#### Results from Sensitivity analyses for primary analyses (using FO+HES cohort)

(i) Excluding those pts who develop neurodegenerative disease or dementia within 10 years after imaging

No additional participants excluded compared to developing neurodegenerative disease or dementia within two years.

(ii) Excluding those pts who had cerebrovascular disease at imaging

96 additional participants excluded from FO+HES cohort when conducted as complete cases for control variables (N=4092; 1:1 RD:control cohorts). Similar to main analyses, 157 (7.7%) in control cohort developed depression after imaging compared to 608 (29.7%) in the RD cohort.

(iii) Using an additional control for those pts with close family history of depression

27 additional participants excluded from FO+HES cohort when conducted as complete cases for control variables (N=4160; 1:1 RD:control cohorts). Similar to main analyses, 140 (6.8%) in control cohort developed depression after imaging compared to 608 (29.7%) in the RD cohort.

(iv) Dividing cohorts into 64 years and less versus 65 and over

In the RD cohort, 1208 participants were 64 years and less, and 886 were 65 and over.

For the <65y: 103 (8.5%) in control cohort developed depression after imaging compared to 371 (30.7%) in the RD cohort

For 65+: 76 (8.6%) in control cohort developed depression after imaging compared to 249 (28.1%) in the RD cohort.

Odds of future depression for control participants in <65: 0.083

Odds of future depression for RD participants in <65: 0.458

Odds of future depression for control participants in >/=65: 0.098

Odds of future depression for RD participants in >/=65: 0.423

Chi squared results between the cohort splits: Xsq = 0.412, p value 0.52

#### Supplementary Table 9: Sensitivity analyses for primary outcome

| **Analyses** |  | **Total n (RD: Control)** |  | **Beta (estimate)** | **Standard error** | **Z score** | **P value** |
| --- | --- | --- | --- | --- | --- | --- | --- |
| ***No neurodegenerative disease or dementia in 10y after imaging*** |  | *4188 (2094: 2094)* |  | Same as main analyses (no additional pts excluded) |  |  |  |
| ***No cerebrovascular disease present at imaging visit*** | *Model without group* | *4092 (2046:2046)* | *Effect of composite cognition score* | 0.0276 | 0.058 | 0.473 | 0.636 |
|  | *Main model* |  | *Effect of composite cognitive score* | -0.296 | 0.135 | -2.621 | 0.0088* |
|  | *Main model* |  | *Composite cognitive score*group* | 0.467 | 0.130 | 3.595 | 0.0003* |
| ***Additionally controlling for close family history of depression*** | *Model without group* | *4188 (2094: 2094)* | *Effect of composite cognition score* | 0.0367 | 0.058 | 0.063 | 0.949 |
|  | *Main model* |  | *Effect of composite cognitive score* | -0.262 | 0.111 | -2.369 | 0.0183* |
|  | *Main model* |  | *Composite cognitive score*group* | 0.409 | 0.128 | 3.198 | 0.0014* |
| ***Only age 64 or less*** | *Main model* | *2416 (1208:1208)* | *Effect of composite cognitive score* | -0.243 | 0.155 | -1.57 | 0.116 |
|  | *Main model* |  | *Composite cognitive score*group* | 0.357 | 0.180 | 1.986 | 0.0471* |
| ***Only age 65 or more*** | *Main model* | *1772 (886: 886)* | *Effect of composite cognitive score* | -0.295 | 0.181 | -1.624 | 0.104 |
|  | *Main model* |  | *Composite cognitive score*group* | 0.556 | 0.211 | 2.627 | 0.0086* |

#### Supplementary Figure 5: Future depression risk relating to composite cognition scores at imaging in the primary cohort: (A) 64 and under subset; (B) 65 and over subset

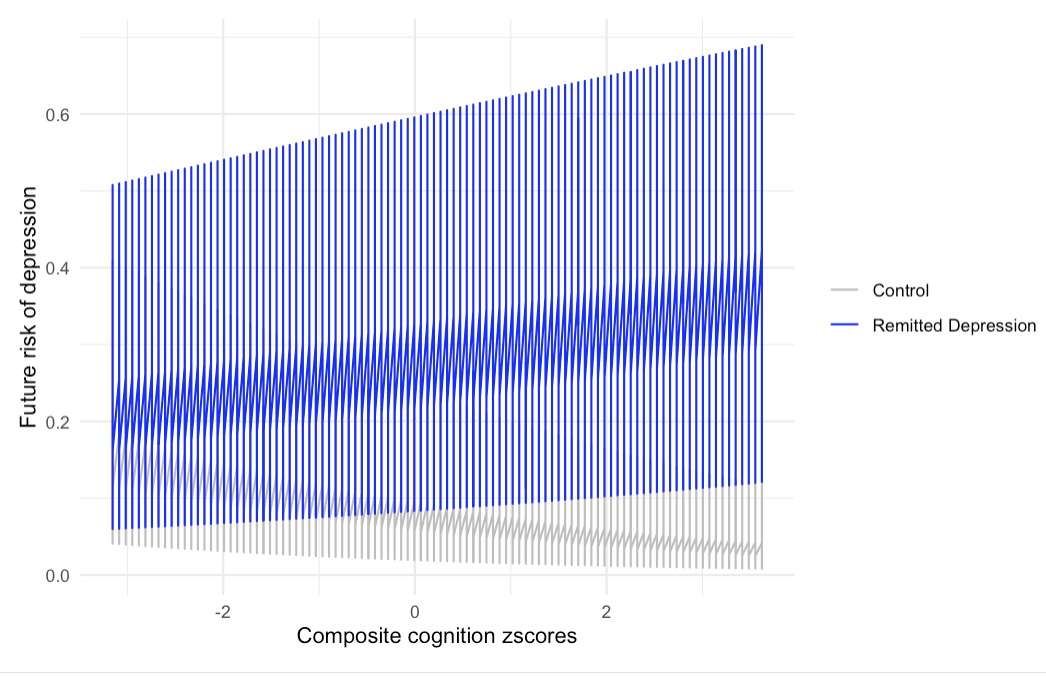

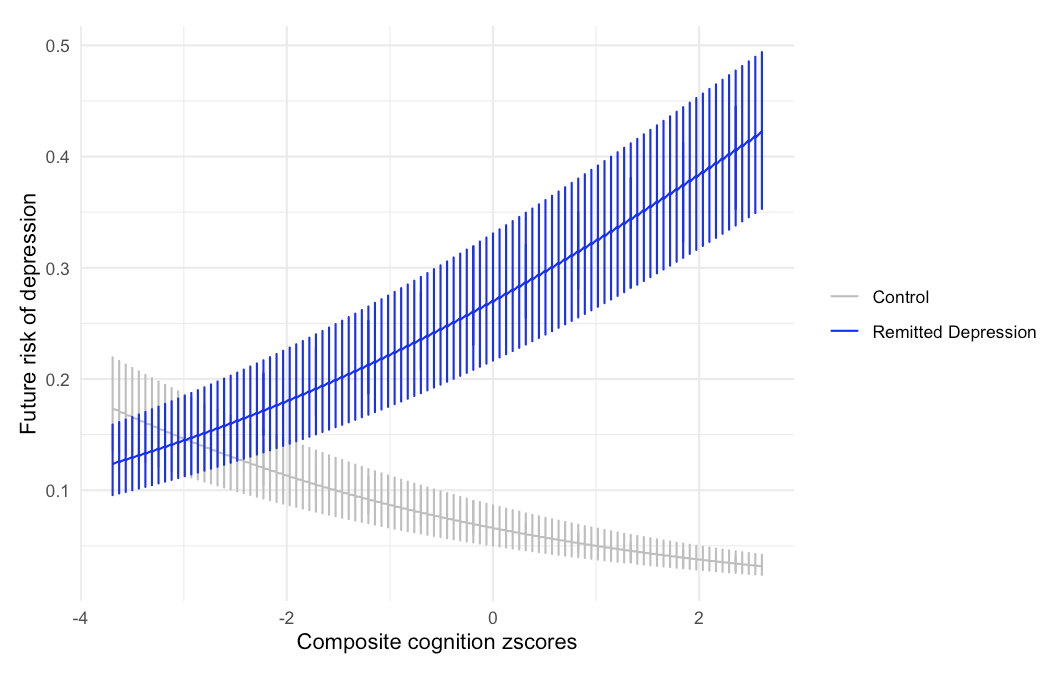

#### Robustness check 1) **Effect on antidepressant use on cognition at imaging within remitted depression group**

#### Supplementary Table 10: Proportion of remitted depression cohort using antidepressants

| **No use** | **Baseline only** | **Baseline & imaging** | **Imaging only** |
| --- | --- | --- | --- |
| 53.01% | 12.56% | 14.33% | 20.11% |

Antidepressant use at baseline and imaging did not appear to have a significant effect on summed cognitive scores at imaging:

i) Comparing no use / antidepressants at baseline only / antidepressants at baseline and imaging / antidepressants only at imaging; p=0.913.
ii) Comparing antidepressants at baseline and imaging versus antidepressants at baseline only (i.e.persistence vs non-persistence); p=0.200

#### Robustness check 2) Overlap between cohorts

#### Supplementary Table 11: Overlap between primary and secondary cohorts

(i) **FO+HES and GP cohort**

| In both subsets | 521 |
| --- | --- |
| Only in FO+HES | 1573 |
| Only in GP | 602 |

(ii) **FO+HES and Probable Depression cohort**

| In both subsets | 385 |
| --- | --- |
| Only in FO+HES | 1709 |
| Only in Probable Depression | 2149 |

(iii) **GP and Probable Depression cohort**

| In both subsets | 254 |
| --- | --- |
| Only in GP | 602 |
| Only in Probable Depression | 2280 |

#### Robustness check 3) **Relapse rate across different confounders**

Out of 2094 participants who fulfilled criteria as defined above for the primary RD cohort (and had full data for control variables), 620 (30%) had a relapse of depression following the UK Biobank visit where cognitive and imaging data were collected. For 2094 matched controls, 179 (8.5%) had a new episode of depression at the equivalent time point.

Of 799 individuals who had an episode of depression after imaging, we can break this down into how this is affected across different non-continuous confounders (%):

#### Supplementary Table 12: Demographics of participants in primary cohort who developed depression after imaging

(i) **Ethnicity**

|  | **White** | **Black** | **Mixed / other** |
| --- | --- | --- | --- |
| ***Controls*** | 96.6 | 1.2 | 2.2 |
| ***RD cohort*** | 99.0 | 0.0 | 1.0 |

(ii) **Education**

|  | **Uni** | **A levels** | **O levels** | **CSEs** | **NVQ/HND** | **Other professional** | **None of the above** |
| --- | --- | --- | --- | --- | --- | --- | --- |
| ***Controls*** | 50.3 | 15.6 | 18.4 | 3.9 | 5.6 | 4.5 | 1.7 |
| ***RD cohort*** | 49.8 | 13.7 | 19.2 | 4.7 | 5.3 | 3.4 | 3.9 |

(iii) **Deprivation**

|  | **Least** |  |  |  | **Most** |
| --- | --- | --- | --- | --- | --- |
| ***Controls*** | 43.0 | 34.6 | 15.6 | 6.7 | 0.0 |
| ***RD cohort*** | 38.7 | 39.2 | 17.7 | 4.2 | 0.2 |

(iv) **Smoking at imaging**

|  | **Non-smoker** | **Smoker** |
| --- | --- | --- |
| ***Controls*** | 52.0 | 48.0 |
| ***RD cohort*** | 54.8 | 45.2 |

(v) **Epilepsy at imaging**

|  | **No** | **Yes** |
| --- | --- | --- |
| ***Controls*** | 98.9 | 1.1 |
| ***RD cohort*** | 99.0 | 1.0 |

(vi) **Chronic IHD at imaging**

|  | **No** | **Yes** |
| --- | --- | --- |
| ***Controls*** | 92.7 | 7.3 |
| ***RD cohort*** | 93.5 | 6.5 |

(vii) **Cerebrovascular disease at imaging**

|  | **No** | **Yes** |
| --- | --- | --- |
| ***Controls*** | 97.2 | 2.8 |
| ***RD cohort*** | 98.1 | 1.9 |

(viii) **Cancer before imaging**

|  | **No** | **Yes** |
| --- | --- | --- |
| ***Controls*** | 86.6 | 13.4 |
| ***RD cohort*** | 85.6 | 14.4 |

#### Supplementary Table 13: Imaging parameters at baseline (n=1554)

| **Parameter** | **Matched control cohort (2094)** | **Primary RD cohort (2094)** | **Estimate** | **P value** |
| --- | --- | --- | --- | --- |
| ***Head motion*** | 0.115 (0.049) | 0.129 (0.064) | 47.4 (F) | **<0.001** |
| ***Total brain volume*** | 1502124 (73011) | 1498992 (73460) | 1.42 (F) | 0.23 |
| ***White matter volume*** | 701931 (40093) | 703397 (39864) | 1.05 (F) | 0.31 |
| ***Grey matter volume*** | 800193 (47085) | 795795 (48417) | 7.20 (F) | **0.007** |
| ***Cortical volume*** | 623404 (40673) | 620398 (41252) | 4.18 (F) | **0.041** |
| ***CSF volume*** | 44305 (18882) | 44837 (18255) | 0.64 (F) | 0.42 |

#### Supplementary Table 14 Composite cognition scores and association with cross-sectional brain regional volumes across all participants (n=1554)

| **Brain region (volume)** | **t value** | **Main effect of region (p value, FDR corrected)** | **Interaction (p value, FDR corrected)** |
| --- | --- | --- | --- |
| *ACC(L)* | -0.04 | 0.97 | 0.92 |
| *ACC(R)* | 0.12 | 0.97 | 0.92 |
| *Amygdala (L)* | 4.63 | **<0.001** | 0.92 |
| *Amygdala (R)* | 5.01 | **<0.001** | 0.92 |
| *Hippocampus (L)* | 3.34 | **0.005** | 0.92 |
| *Hippocampus (R)* | 2.77 | **0.02** | 0.92 |
| *Middle frontal gyrus (L)* | 2.28 | **0.049** | 0.96 |
| *Middle frontal gyrus (R)* | 2.25 | **0.049** | 0.92 |
| *Precuneous (L)* | 2.58 | **0.03** | 0.93 |
| *Precuneous (R)* | 1.91 | 0.09 | 0.96 |
| *PCC(L)* | 2.44 | **0.039** | 0.96 |
| *PCC(R)* | 2.14 | 0.05 | 0.96 |
| *Supramarginal gyrus (L)* | 0.05 | 0.97 | 0.92 |
| *Supramarginal gyrus (R)* | 1.00 | 0.39 | 0.92 |
| *Superior frontal gyrus (L)* | 1.22 | 0.33 | 0.92 |
| *Superior frontal gyrus (R)* | 1.02 | 0.34 | 0.92 |

#### Supplementary Table 15 (a): Regional structural volumes of the Triple Network for RD cohort with full imaging data (Group 1; n=1554) and matched control cohort (Group 0; n=1554)

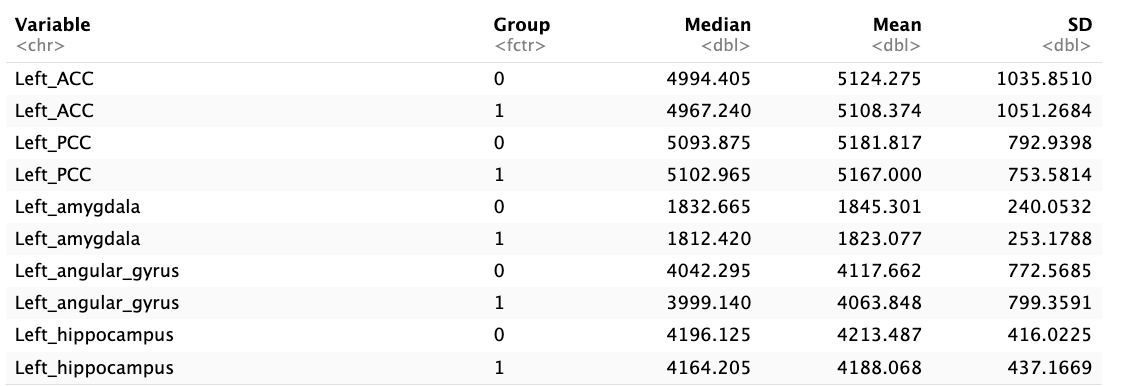

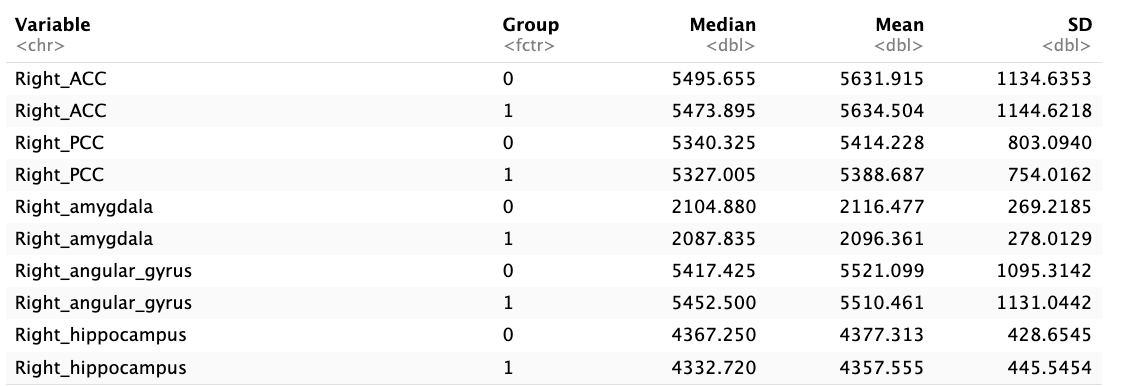

#### Supplementary Table 15 (b): Group differences in structural imaging for RD cohort with full imaging data (n=1554) and matched control cohort (n=1554)

*Controlled for total brain volume and age and sex orthogonally as well as remainder of pre-specified covariates, and also shown with adjustment for FDR*

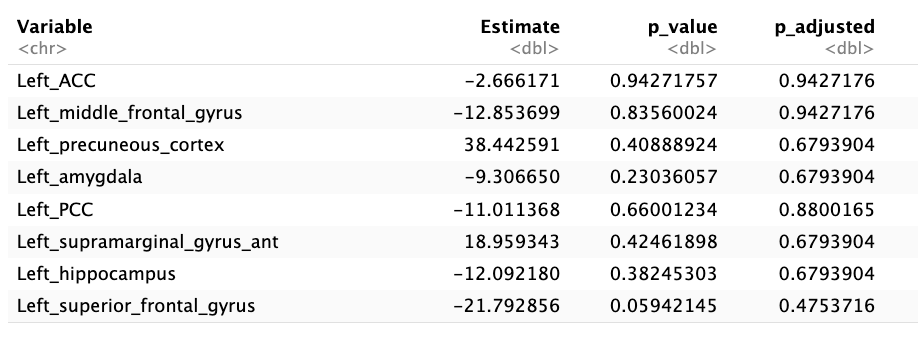

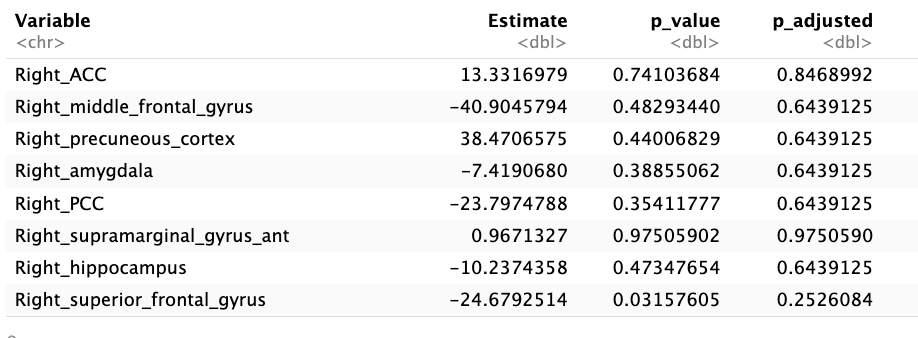

#### Supplementary Table 16: Impact of brain regional volumes within Triple Network on future risk of depression for RD (n=1554) and matched control cohorts (n=1554)

(i) Main effect of group

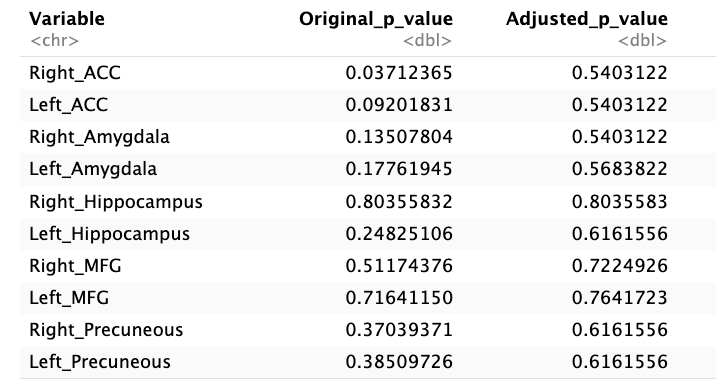

(ii) Group*regional volume interaction

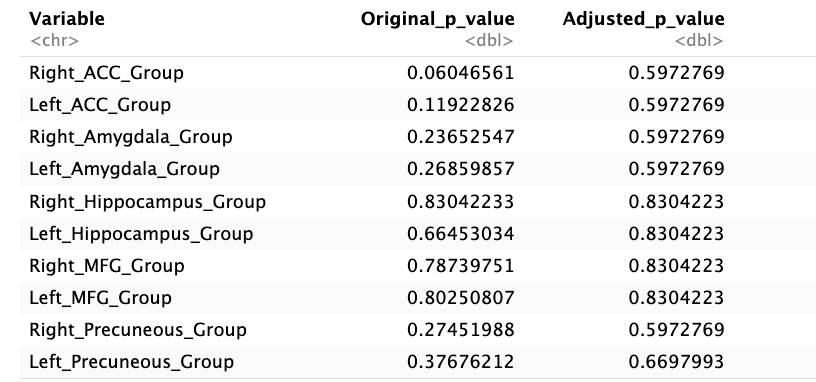

#### Supplementary Table 17: Mediation analyses of significant structural results (controlled for potential confounders)

(i) Left PCC

| Cognition | Direct effect (estimate, p value) | Indirect effect (estimate, p value) | Total effect (estimate, p value) |
| --- | --- | --- | --- |
| *Composite cognition* | 7.52 x 10^-6^; p=0.31 | 3.11 x 10^-7^; p=0.81 | 7.83 x 10^-6^; p=0.27 |
| *Memory* | -7.26 x 10^-6^; p=0.64 | -4.96 x 10^-8^; p=0.94 | -7.31 x 10^-6^ ; p=0.64 |
| *Executive function* | 7.80 x 10^-6^; p=0.29 | 7.61 x 10^-7^; p=0.49 | 8.56 x 10^-6^ ; p=0.23 |
| *Reasoning* | 7.80 x 10^-6^; p=0.28 | -3.68 x 10^-7^; p=0.84 | 7.45 x 10^-6^ ; p=0.30 |
| *Processing speed* | 7.25 x 10^-6^; p=0.33 | -4.98 x 10^-7^; p=0.84 | 6.75 x 10^-6^ ; p=0.35 |

(ii) Right PCC

| Cognition | Direct effect (estimate, p value) | Indirect effect (estimate, p value) | Total effect (estimate, p value) |
| --- | --- | --- | --- |
| *Composite cognition* | 7.72 x 10^-6^; p=0.31 | 3.34 x 10^-7^; p=0.85 | 7.38 x 10^-6^; p=0.31 |
| *Memory* | -2.04 x 10^-6^; p=0.99 | -2.18 x 10^-8^; p=0.92 | -2.06 x 10^-6^ ; p=0.98 |
| *Executive function* | 8.50 x 10^-6^; p=0.21 | 7.21 x 10^-7^; p=0.50 | 9.22 x 10^-6^ ; p=0.18 |
| *Reasoning* | 8.95 x 10^-6^; p=0.21 | -3.96 x 10^-7^; p=0.82 | 8.56 x 10^-6^ ; p=0.22 |
| *Processing speed* | 8.40 x 10^-6^; p=0.25 | -3.99 x 10^-7^; p=0.70 | 8.01 x 10^-6^ ; p=0.27 |

(iii) Left amygdala

| Cognition | Direct effect (estimate, p value) | Indirect effect (estimate, p value) | Total effect (estimate, p value) |
| --- | --- | --- | --- |
| *Composite cognition* | -6.09 x 10^-5^; p=0.27 | 5.77 x 10^-6^; p=0.65 | -5.51 x 10^-5^; p=0.34 |
| *Memory* | -4.40 x 10^-5^; p=0.33 | -1.40 x 10^-7^; p=0.91 | -4.41 x 10^-5^ ; p=0.32 |
| *Executive function* | -6.07 x 10^-5^; p=0.28 | 7.71 x 10^-6^; p=0.43 | -5.30 x 10^-5^; p=0.38 |
| *Reasoning* | -6.96 x 10^-5^; p=0.22 | 4.53 x 10^-8^; p=0.97 | -6.96 x 10^-5^ ; p=0.22 |
| *Processing speed* | -7.31 x 10^-5^; p=0.19 | -3.19 x 10^-6^; p=0.77 | -7.62 x 10^-5^ ; p=0.17 |

(iv) Right amygdala

| Cognition | Direct effect (estimate, p value) | Indirect effect (estimate, p value) | Total effect (estimate, p value) |
| --- | --- | --- | --- |
| *Composite cognition* | -8.24 x 10^-5^; p=0.05 | 1.77 x 10^-5^; p=0.180 | -4.41 x 10^-5^ ; p=0.32 |
| *Memory* | -4.30 x 10^-5^; p=0.31 | -2.10 x 10^-7^; p=0.85 | -4.32 x 10^-5^ ; p=0.31 |
| *Executive function* | -7.39 x 10^-5^; p=0.16 | 7.83 x 10^-6^; p=0.37 | -6.60 x 10^-5^; p=0.22 |
| *Reasoning* | -7.70 x 10^-5^; p=0.15 | -4.02 x 10^-7^; p=0.98 | -7.74 x 10^-5^ ; p=0.14 |
| *Processing speed* | -7.77 x 10^-5^; p=0.14 | -2.86 x 10^-6^; p=0.78 | -8.06 x 10^-5^ ; p=0.12 |

(v) Left ACC

| Cognition | Direct effect (estimate, p value) | Indirect effect (estimate, p value) | Total effect (estimate, p value) |
| --- | --- | --- | --- |
| *Composite cognition* | 9.15 x 10^-7^; p=0.81 | 1.32 x 10^-7^; p=0.77 | 1.05 x 10^-6^ ; p=0.81 |
| *Memory* | -6.99 x 10^-6^; p=0.49 | 8.45 x 10^-8^; p=0.80 | -6.91 x 10^-6^ ; p=0.49 |
| *Executive function* | 1.85 x 10^-6^; p=0.69 | 2.49 x 10^-7^; p=0.50 | 2.10 x 10^-6^; p=0.67 |
| *Reasoning* | 2.15 x 10^-6^; p=0.66 | -9.54 x 10^-8^; p=0.92 | 2.05 x 10^-6^ ; p=0.68 |
| *Processing speed* | 2.84 x 10^-6^; p=0.58 | -6.17 x 10^-8^; p=0.83 | 2.77 x 10^-6^ ; p=0.58 |

(vi) Right ACC

| Cognition | Direct effect (estimate, p value) | Indirect effect (estimate, p value) | Total effect (estimate, p value) |
| --- | --- | --- | --- |
| *Composite cognition* | -3.97 x 10^-7^; p=0.92 | 1.24 x 10^-7^; p=0.74 | -2.73 x 10^-7^ ; p=0.92 |
| *Memory* | -7.90 x 10^-6^; p=0.34 | 5.99 x 10^-8^; p=0.83 | -7.84 x 10^-6^ ; p=0.34 |
| *Executive function* | 4.35 x 10^-7^; p=0.81 | 1.77 x 10^-7^; p=0.53 | 6.12 x 10^-7^; p=0.81 |
| *Reasoning* | 3.14 x 10^-7^; p=0.86 | -8.81 x 10^-8^; p=0.91 | 2.26 x 10^-7^ ; p=0.87 |
| *Processing speed* | 2.09 x 10^-6^; p=0.66 | -8.00 x 10^-8^; p=0.73 | 2.01 x 10^-6^ ; p=0.67 |

(vii) Left hippocampus

| Cognition | Direct effect (estimate, p value) | Indirect effect (estimate, p value) | Total effect (estimate, p value) |
| --- | --- | --- | --- |
| *Composite cognition* | -1.86 x 10^-5^; p=0.60 | 1.82 x 10^-6^; p=0.72 | -1.68 x 10^-5^ ; p=0.65 |
| *Memory* | -4.19 x 10^-5^; p=0.13 | -1.34 x 10^-7^; p=0.86 | -4.21 x 10^-5^ ; p=0.13 |
| *Executive function* | -1.89 x 10^-5^; p=0.60 | 3.08 x 10^-6^; p=0.42 | -1.58 x 10^-5^; p=0.70 |
| *Reasoning* | -2.21 x 10^-5^; p=0.55 | -4.96 x 10^-7^; p=0.94 | -2.26 x 10^-5^ ; p=0.52 |
| *Processing speed* | -2.31 x 10^-5^; p=0.52 | -1.65 x 10^-6^; p=0.79 | -2.47 x 10^-5^ ; p=0.47 |

(viii) Right hippocampus

| Cognition | Direct effect (estimate, p value) | Indirect effect (estimate, p value) | Total effect (estimate, p value) |
| --- | --- | --- | --- |
| *Composite cognition* | 4.16 x 10^6^; p=0.58 | 9.83 x 10^7^; p=0.78 | 5.14 x 10^6^ ; p=0.51 |
| *Memory* | -1.18 x 10^5^; p=0.71 | -9.36 x 10^8^; p=0.89 | -1.19 x 10^5^ ; p=0.71 |
| *Executive function* | 4.15 x 10^6^; p=0.56 | 2.00 x 10^6^; p=0.40 | 6.15 x 10^6^; p=0.48 |
| *Reasoning* | 4.83 x 10^6^; p=0.54 | -7.03 x 10^7^; p=0.82 | 4.12 x 10^6^ ; p=0.57 |
| *Processing speed* | 4.25 x 10^6^; p=0.58 | -1.23 x 10^6^; p=0.75 | 3.02 x 10^6^ ; p=0.62 |

(ix) Left precuneous cortex

| Cognition | Direct effect (estimate, p value) | Indirect effect (estimate, p value) | Total effect (estimate, p value) |
| --- | --- | --- | --- |
| *Composite cognition* | 1.73 x 10^-6^; p=0.60 | 2.95 x 10^-7^; p=0.81 | 2.03 x 10^-6^; p=0.56 |
| *Memory* | -4.01 x 10^-7^; p=0.90 | -3.35 x 10^-8^; p=0.86 | -4.35 x 10^-7^ ; p=0.90 |
| *Executive function* | 1.15 x 10^-6^; p=0.68 | 6.61 x 10^-7^; p=0.47 | 1.81 x 10^-6^; p=0.58 |
| *Reasoning* | 1.46 x 10^-6^; p=0.65 | -1.78 x 10^-7^; p=0.88 | 1.28 x 10^-6^ ; p=0.67 |
| *Processing speed* | 1.54 x 10^-6^; p=0.65 | -4.54 x 10^-7^; p=0.73 | 1.08 x 10^-6^ ; p=0.70 |

(x) Right precuneous cortex

| Cognition | Direct effect (estimate, p value) | Indirect effect (estimate, p value) | Total effect (estimate, p value) |
| --- | --- | --- | --- |
| *Composite cognition* | 3.35 x 10^-6^; p=0.34 | 2.62 x 10^-7^; p=0.77 | 3.62 x 10^-6^; p=0.31 |
| *Memory* | 9.10 x 10^-7^; p=0.70 | -1.57 x 10^-8^; p=0.88 | 8.94 x 10^-7^ ; p=0.70 |
| *Executive function* | 2.85 x 10^-6^; p=0.44 | 5.21 x 10^-7^; p=0.47 | 3.37 x 10^-6^; p=0.38 |
| *Reasoning* | 3.50 x 10^-6^; p=0.34 | -1.83 x 10^-7^; p=0.86 | 3.32 x 10^-6^ ; p=0.35 |
| *Processing speed* | 3.29 x 10^-6^; p=0.37 | -4.24 x 10^-7^; p=0.68 | 2.86 x 10^-6^ ; p=0.41 |

(xi) Left middle frontal gyrus

| Cognition | Direct effect (estimate, p value) | Indirect effect (estimate, p value) | Total effect (estimate, p value) |
| --- | --- | --- | --- |
| *Composite cognition* | -6.79 x 10^-7^; p=0.99 | 2.90 x 10^-7^; p=0.77 | -3.89 x 10^-7^; p=0.93 |
| *Memory* | -1.83 x 10^-6^; p=0.81 | -1.44 x 10^-8^; p=0.93 | -1.84 x 10^-6^ ; p=0.81 |
| *Executive function* | -1.22 x 10^-6^; p=0.91 | 5.08 x 10^-7^; p=0.42 | -7.14 x 10^-7^; p=1.00 |
| *Reasoning* | -9.95 x 10^-7^; p=0.99 | -1.08 x 10^-7^; p=0.91 | -1.10 x 10^-6^ ; p=0.95 |
| *Processing speed* | -4.21 x 10^-7^; p=0.95 | -3.52 x 10^-7^; p=0.67 | -7.73 x 10^-7^ ; p=0.98 |

(xii) Right middle frontal gyrus

| Cognition | Direct effect (estimate, p value) | Indirect effect (estimate, p value) | Total effect (estimate, p value) |
| --- | --- | --- | --- |
| *Composite cognition* | 6.44 x 10^-7^; p=0.96 | 2.94 x 10^-7^; p=0.77 | 9.38 x 10^-7^; p=0.90 |
| *Memory* | -5.06 x 10^-6^; p=0.39 | -1.93 x 10^-9^; p=0.99 | -5.06 x 10^-6^ ; p=0.38 |
| *Executive function* | -4.62 x 10^-7^; p=0.91 | 5.27 x 10^-7^; p=0.48 | 6.49 x 10^-8^; p=0.85 |
| *Reasoning* | 1.62 x 10^-7^; p=0.81 | -8.37 x 10^-8^; p=0.93 | 7.83 x 10^-8^; p=0.83 |
| *Processing speed* | -1.83 x 10^-8^; p=0.88 | -3.34 x 10^-7^; p=0.71 | -3.52 x 10^-7^ ; p=0.92 |

#### Supplementary Table 18 (a): Impact of rsfMRI functional connectivity (nodes) on cognitive scores within Triple Network

|  | Main effect | | | Node*group | | |
| --- | --- | --- | --- | --- | --- | --- |
| Node | Estimate | Unadjusted p values | P values with FDR correction | Estimate | Unadjusted p values | P values with FDR correction |
| X1 (DMN) | *-0.016* | *0.81* | *0.9470* | *-0.044* | *0.65* | *0.8965* |
| X7 (DMN / cEN) | *0.0051* | *0.95* | *0.9470* | *-0.0002* | *0.99* | *0.9983* |
| X9 (DMN / cEN) | *0.029* | *0.64* | *0.9470* | *-0.014* | *0.87* | *0.9564* |
| X14 (DMN / cEN) | *0.046* | *0.68* | *0.9470* | *-0.083* | *0.60* | *0.8965* |
| X20 (DMN / cEN) | *-0.055* | *0.50* | *0.9470* | *0.131* | *0.27* | *0.8965* |
| X5 (cEN / DMN) | *0.083* | *0.26* | *0.9470* | *-0.038* | *0.73* | *0.8965* |
| X6 (cEN / DMN) | *0.068* | *0.40* | *0.9470* | *-0.053* | *0.66* | *0.8965* |
| X16 (cEN) | *-0.0088* | *0.91* | *0.9470* | *-0.055* | *0.64* | *0.8965* |
| X21 (cEN / DMN) | *0.071* | *0.43* | *0.9470* | *-0.066* | *0.61* | *0.8965* |
| X3 (SN) | *-0.016* | *0.84* | *0.9470* | *0.052* | *0.65* | *0.8965* |
| X13 (SN / DMN) | *-0.164* | *0.08* | *0.8964* | *0.118* | *0.37* | *0.8965* |

#### Supplementary Table 18 (b): Impact of rsfMRI functional connectivity (edges) on cognitive scores within Triple Network

|  | **Main effect** | | | **Edge*group** | | |
| --- | --- | --- | --- | --- | --- | --- |
| **Edge** | **Estimate** | **Unadjusted p values** | **P values with FDR correction** | **Estimate** | Unadjusted p values | **P values with FDR correction** |
| ***V2 (nodes 1-3) DMN - SN*** | 0.0056 | 0.8534 | 0.9839 | 0.0168 | 0.6923 | 0.8862 |
| ***V7 (nodes 1-5) DMN – cEN / DMN)*** | 0.0146 | 0.6273 | 0.9137 | -0.0183 | 0.6753 | 0.8862 |
| ***V9 (nodes 3-5) SN – cEN / DMN*** | -0.0392 | 0.1853 | 0.9137 | 0.0269 | 0.5241 | 0.8479 |
| ***V11 (nodes 1-6) DMN – cEN / DMN*** | 0.0212 | 0.5218 | 0.9137 | 0.0309 | 0.5180 | 0.8479 |
| ***V13 (nodes 3-6) SN – cEN / DMN*** | -0.0045 | 0.8895 | 0.9839 | 0.0050 | 0.9142 | 0.9669 |
| ***V15 (nodes 5-6) cEN / DMN – cEN / DMN*** | -0.0147 | 0.6136 | 0.9137 | -0.0132 | 0.7504 | 0.8920 |
| ***V16 (nodes 1-7) DMN – DMN / cEN*** | -0.0013 | 0.9674 | 0.9839 | 0.0016 | 0.9713 | 0.9893 |
| ***V18 (nodes 3-7) SN – DMN / cEN*** | 0.0364 | 0.2323 | 0.9137 | -0.0493 | 0.2500 | 0.7411 |
| ***V20 (nodes 5-7) cEN / DMN – DMN / cEN*** | 0.0055 | 0.8554 | 0.9839 | -0.0468 | 0.2830 | 0.7411 |
| ***V21 (nodes 6-7) cEN / DMN – DMN / cEN*** | -0.0007 | 0.9839 | 0.9839 | -0.0529 | 0.2586 | 0.7411 |
| ***V29 (nodes 1-9) DMN – DMN / cEN*** | 0.0415 | 0.1869 | 0.9137 | -0.0431 | 0.3385 | 0.8066 |
| ***V31 (nodes 3-9) SN – DMN / cEN*** | 0.0109 | 0.7426 | 0.9839 | -0.0006 | 0.9893 | 0.9893 |
| ***V33 (nodes 5-9) cEN / DMN – DMN / cEN*** | -0.0373 | 0.2012 | 0.9137 | 0.0747 | 0.0807 | 0.7411 |
| ***V34 (nodes 6-9) cEN / DMN – DMN / cEN*** | -0.0183 | 0.5641 | 0.9137 | 0.0190 | 0.6719 | 0.8862 |
| ***V35 (nodes 7-9) DMN / cEN - cEN*** | 0.0171 | 0.5857 | 0.9137 | -0.0559 | 0.2072 | 0.7411 |
| ***V67 (nodes 1-13) DMN – SN / DMN*** | 0.0025 | 0.9473 | 0.9839 | -0.0717 | 0.1829 | 0.7411 |
| ***V69 (nodes 3-13) SN – DMN*** | 0.0546 | 0.1164 | 0.9137 | -0.1536 | 0.0027 | 0.1500 |
| ***V71 (nodes 5-13) cEN / DMN – DMN*** | 0.0152 | 0.6258 | 0.9137 | -0.0265 | 0.5442 | 0.8552 |
| ***V72 (nodes 6-13) cEN / DMN – DMN*** | -0.0160 | 0.6313 | 0.9137 | -0.0338 | 0.4888 | 0.8402 |
| ***V73 (nodes 7-13) DMN / cEN – DMN*** | 0.0010 | 0.9785 | 0.9839 | 0.0576 | 0.2782 | 0.7411 |
| ***V75 (nodes 9-13) cEN – DMN*** | -0.0308 | 0.3699 | 0.9137 | 0.0668 | 0.1828 | 0.7411 |
| ***V79 (nodes 1-14) DMN – DMN / cEN*** | 0.0310 | 0.3234 | 0.9137 | -0.0710 | 0.1211 | 0.7411 |
| ***V81 (nodes 3-14) SN – DMN / cEN*** | -0.0253 | 0.5386 | 0.9137 | 0.0079 | 0.8931 | 0.9631 |
| ***V83 (nodes 5-14) cEN / DMN – DMN / cEN*** | -0.0075 | 0.8481 | 0.9839 | -0.0831 | 0.1324 | 0.7411 |
| ***V84 (nodes 6-14) cEN / DMN – DMN / cEN*** | 0.0435 | 0.2868 | 0.9137 | -0.0969 | 0.0880 | 0.7411 |
| ***V85 (nodes 7-14) DMN / cEN – DMN / cEN*** | 0.0246 | 0.5193 | 0.9137 | -0.0149 | 0.7873 | 0.8920 |
| ***V87 (nodes 9-14) cEN – DMN / cEN*** | -0.0308 | 0.4293 | 0.9137 | 0.0948 | 0.0994 | 0.7411 |
| ***V91 (nodes 13-14) DMN – DMN / cEN*** | -0.0400 | 0.3279 | 0.9137 | 0.0749 | 0.2180 | 0.7411 |
| ***V106 (nodes 1-16) DMN – cEN*** | 0.0142 | 0.6126 | 0.9137 | 0.0355 | 0.3666 | 0.8066 |
| ***V108 (nodes 3-16) SN – cEN*** | 0.0297 | 0.3480 | 0.9137 | -0.0558 | 0.2263 | 0.7411 |
| ***V110 (nodes 5-16) cEN / DMN – cEN*** | 0.0349 | 0.2316 | 0.9137 | -0.0138 | 0.7470 | 0.8920 |
| ***V111 (nodes 6-16) cEN / DMN – cEN*** | 0.0027 | 0.9282 | 0.9839 | -0.0424 | 0.3112 | 0.7779 |
| ***V112 (nodes 7-16) DMN / cEN – cEN*** | -0.0168 | 0.5646 | 0.9137 | 0.0542 | 0.1963 | 0.7411 |
| ***V114 (nodes 9-16) cEN – cEN*** | 0.0278 | 0.3789 | 0.9137 | -0.0241 | 0.5872 | 0.8729 |
| ***V118 (nodes 13-16) DMN – cEN*** | -0.0438 | 0.2326 | 0.9137 | 0.0296 | 0.5736 | 0.8729 |
| ***V119 (nodes 14 – 16) cEN – cEN*** | 0.0445 | 0.2223 | 0.9137 | -0.0361 | 0.4877 | 0.8402 |
| ***V172 (nodes 1-20) DMN – DMN / cEN*** | 0.0212 | 0.4857 | 0.9137 | 0.0173 | 0.6929 | 0.8862 |
| ***V174 (nodes 3-20) SN – DMN / CEN*** | -0.0177 | 0.6058 | 0.9137 | -0.0142 | 0.7725 | 0.8920 |
| ***V176 (nodes 5-20) cEN / DMN – DMN / cEN*** | -0.0479 | 0.1265 | 0.9137 | 0.0363 | 0.4328 | 0.8330 |
| ***V177 (nodes 6-20) cEN – DMN / cEN*** | 0.0673 | 0.0280 | 0.9137 | -0.0651 | 0.1423 | 0.7411 |
| ***V178 (nodes 7-20) DMN / cEN – DMN / cEN*** | 0.0241 | 0.4096 | 0.9137 | -0.0350 | 0.4105 | 0.8330 |
| ***V180 (nodes 9-20) cEN – DMN / cEN*** | -0.0164 | 0.6245 | 0.9137 | -0.0206 | 0.6740 | 0.8862 |
| ***V184 (nodes 13-20) DMN – DMN / cEN*** | -0.0599 | 0.1166 | 0.9137 | -0.0184 | 0.7356 | 0.8920 |
| ***V185 (nodes 14-20) cEN – DMN / cEN*** | -0.0069 | 0.8520 | 0.9839 | -0.0045 | 0.9332 | 0.9685 |
| ***V191 (nodes 1-21) DMN – cEN / DMN*** | -0.0131 | 0.6934 | 0.9137 | 0.0799 | 0.1042 | 0.7411 |
| ***V193 (nodes 3-21) SN – cEN / DMN*** | -0.0085 | 0.8129 | 0.9839 | 0.0424 | 0.4133 | 0.8330 |
| ***V195 (nodes 5-21) cEN / DMN – cEN / DMN*** | -0.0052 | 0.8657 | 0.9839 | 0.0112 | 0.7947 | 0.8920 |
| ***V196 (nodes 6-21) cEN – cEN / DMN*** | -0.0231 | 0.3760 | 0.9137 | 0.0263 | 0.4848 | 0.8402 |
| ***V197 (nodes 7-21) DMN / cEN – cEN / DMN*** | -0.0674 | 0.0510 | 0.9137 | 0.0723 | 0.1456 | 0.7411 |
| ***V199 (nodes 9-21) cEN – cEN / DMN*** | -0.0117 | 0.7319 | 0.9839 | 0.0535 | 0.2795 | 0.7411 |
| ***V203 (nodes 13-21) DMN – cEN / DMN*** | -0.0716 | 0.0488 | 0.9137 | 0.0865 | 0.0973 | 0.7411 |
| ***V204 (nodes 14-21) cEN – cEN / DMN*** | 0.0011 | 0.9773 | 0.9839 | 0.0445 | 0.4392 | 0.8330 |
| ***V206 (nodes 16-21) cEN – cEN / DMN*** | -0.0213 | 0.5021 | 0.9137 | 0.0411 | 0.3645 | 0.8066 |
| ***V210 (nodes 20-21) DMN / cEN – cEN / DMN*** | -0.0189 | 0.5886 | 0.9137 | 0.0241 | 0.6425 | 0.8862 |

#### Supplementary Table 19 (a): Impact of rsfMRI functional connectivity (nodes) on future risk of depression

|  | **Main effect** | | | **Node*group** | | |
| --- | --- | --- | --- | --- | --- | --- |
| **Node** | **Estimate** | **Unadjusted p values** | **P values with FDR correction** | **Estimate** | **Unadjusted p values** | **P values with FDR correction** |
| ***X1 (DMN)*** | 0.0050 | 0.8464 | 0.8550 | -0.0316 | 0.3949 | 0.9830 |
| ***X7 (DMN / cEN)*** | -0.0078 | 0.7979 | 0.8550 | 0.0034 | 0.9386 | 0.9830 |
| ***X9 (DMN / cEN)*** | 0.0424 | 0.1390 | 0.8550 | -0.0296 | 0.4697 | 0.9830 |
| ***X14 (DMN / cEN)*** | 0.0144 | 0.6439 | 0.8550 | -0.0151 | 0.7373 | 0.9830 |
| ***X20 (DMN / cEN)*** | 0.0134 | 0.6565 | 0.8550 | 0.0009 | 0.9830 | 0.9830 |
| ***X5 (cEN / DMN)*** | -0.0101 | 0.6667 | 0.8550 | 0.0434 | 0.1901 | 0.9830 |
| ***X6 (cEN / DMN)*** | -0.0068 | 0.8499 | 0.8550 | 0.0106 | 0.8333 | 0.9830 |
| ***X16 (cEN)*** | -0.0092 | 0.8245 | 0.8550 | 0.0255 | 0.6726 | 0.9830 |
| ***X21 (cEN / DMN)*** | -0.0064 | 0.8321 | 0.8550 | 0.0478 | 0.2701 | 0.9830 |
| ***X3 (SN)*** | -0.0057 | 0.8550 | 0.8550 | 0.0109 | 0.8045 | 0.9830 |
| ***X13 (SN / DMN)*** | -0.0396 | 0.2566 | 0.8550 | -0.0415 | 0.4017 | 0.9830 |

#### Supplementary Table 19 (b): Impact of rsfMRI functional connectivity (edges) on future risk of depression

|  | **Main effect** | | | **Edge*group** | | |
| --- | --- | --- | --- | --- | --- | --- |
| **Edge** | **Estimate** | **Unadjusted p values** | **P values with FDR correction** | **Estimate** | **Unadjusted p values** | **P values with FDR correction** |
| ***V2 (nodes 1-3) DMN - SN*** | 0.0062 | 0.5887 | 0.9862 | 0.0120 | 0.4473 | 0.8884 |
| ***V7 (nodes 1-5) DMN – cEN / DMN)*** | 0.0021 | 0.8569 | 0.9862 | 0.0282 | 0.0848 | 0.6769 |
| ***V9 (nodes 3-5) SN – cEN / DMN*** | -0.0124 | 0.2703 | 0.9862 | 0.0015 | 0.9272 | 0.9582 |
| ***V11 (nodes 1-6) DMN – cEN / DMN*** | 0.0015 | 0.9028 | 0.9862 | -0.0113 | 0.5295 | 0.9155 |
| ***V13 (nodes 3-6) SN – cEN / DMN*** | -0.0020 | 0.8706 | 0.9862 | -0.0075 | 0.6682 | 0.9232 |
| ***V15 (nodes 5-6) cEN / DMN – cEN / DMN*** | 0.0002 | 0.9862 | 0.9862 | 0.0145 | 0.3510 | 0.8884 |
| ***V16 (nodes 1-7) DMN – DMN / cEN*** | -0.0087 | 0.4685 | 0.9862 | 0.0122 | 0.4704 | 0.8921 |
| ***V18 (nodes 3-7) SN – DMN / cEN*** | -0.0012 | 0.9160 | 0.9862 | 0.0058 | 0.7218 | 0.9232 |
| ***V20 (nodes 5-7) cEN / DMN – DMN / cEN*** | -0.0048 | 0.6767 | 0.9862 | -0.0012 | 0.9408 | 0.9582 |
| ***V21 (nodes 6-7) cEN / DMN – DMN / cEN*** | 0.0007 | 0.9542 | 0.9862 | 0.0017 | 0.9242 | 0.9582 |
| ***V29 (nodes 1-9) DMN – DMN / cEN*** | 0.0072 | 0.5446 | 0.9862 | -0.0005 | 0.9777 | 0.9777 |
| ***V31 (nodes 3-9) SN – DMN / cEN*** | 0.0103 | 0.4061 | 0.9862 | -0.0065 | 0.7113 | 0.9232 |
| ***V33 (nodes 5-9) cEN / DMN – DMN / cEN*** | -0.0020 | 0.8556 | 0.9862 | 0.0090 | 0.5754 | 0.9232 |
| ***V34 (nodes 6-9) cEN / DMN – DMN / cEN*** | 0.0079 | 0.5074 | 0.9862 | 0.0102 | 0.5493 | 0.9155 |
| ***V35 (nodes 7-9) DMN / cEN - cEN*** | -0.0080 | 0.4959 | 0.9862 | 0.0212 | 0.1992 | 0.6847 |
| ***V67 (nodes 1-13) DMN – SN / DMN*** | 0.0117 | 0.4192 | 0.9862 | -0.0271 | 0.1824 | 0.6769 |
| ***V69 (nodes 3-13) SN – DMN*** | -0.0041 | 0.7625 | 0.9862 | -0.0205 | 0.2936 | 0.8884 |
| ***V71 (nodes 5-13) cEN / DMN – DMN*** | -0.0012 | 0.9161 | 0.9862 | -0.0082 | 0.6170 | 0.9232 |
| ***V72 (nodes 6-13) cEN / DMN – DMN*** | 0.0043 | 0.7387 | 0.9862 | 0.0119 | 0.5137 | 0.9155 |
| ***V73 (nodes 7-13) DMN / cEN – DMN*** | -0.0030 | 0.8326 | 0.9862 | 0.0229 | 0.2535 | 0.8201 |
| ***V75 (nodes 9-13) cEN – DMN*** | 0.0014 | 0.9176 | 0.9862 | -0.0076 | 0.6902 | 0.9232 |
| ***V79 (nodes 1-14) DMN – DMN / cEN*** | 0.0068 | 0.5669 | 0.9862 | -0.0137 | 0.4269 | 0.8884 |
| ***V81 (nodes 3-14) SN – DMN / cEN*** | 0.0068 | 0.6632 | 0.9862 | -0.0217 | 0.3242 | 0.8884 |
| ***V83 (nodes 5-14) cEN / DMN – DMN / cEN*** | -0.0112 | 0.4494 | 0.9862 | -0.0041 | 0.8427 | 0.9560 |
| ***V84 (nodes 6-14) cEN / DMN – DMN / cEN*** | 0.0179 | 0.2472 | 0.9862 | -0.0165 | 0.4414 | 0.8884 |
| ***V85 (nodes 7-14) DMN / cEN – DMN / cEN*** | 0.0066 | 0.6510 | 0.9862 | -0.0293 | 0.1572 | 0.6769 |
| ***V87 (nodes 9-14) cEN – DMN / cEN*** | -0.0110 | 0.4522 | 0.9862 | 0.0092 | 0.6652 | 0.9232 |
| ***V91 (nodes 13-14) DMN – DMN / cEN*** | 0.0126 | 0.4188 | 0.9862 | -0.0315 | 0.1667 | 0.6769 |
| ***V106 (nodes 1-16) DMN – cEN*** | -0.0073 | 0.4905 | 0.9862 | 0.0061 | 0.6830 | 0.9232 |
| ***V108 (nodes 3-16) SN – cEN*** | 0.0163 | 0.1701 | 0.9862 | -0.0273 | 0.1079 | 0.6769 |
| ***V110 (nodes 5-16) cEN / DMN – cEN*** | 0.0041 | 0.7126 | 0.9862 | 0.0098 | 0.5459 | 0.9155 |
| ***V111 (nodes 6-16) cEN / DMN – cEN*** | -0.0124 | 0.2669 | 0.9862 | -0.0235 | 0.1307 | 0.6769 |
| ***V112 (nodes 7-16) DMN / cEN – cEN*** | -0.0109 | 0.3288 | 0.9862 | -0.0019 | 0.9040 | 0.9582 |
| ***V114 (nodes 9-16) cEN – cEN*** | 0.0080 | 0.5030 | 0.9862 | 0.0033 | 0.8419 | 0.9560 |
| ***V118 (nodes 13-16) DMN – cEN*** | -0.0035 | 0.8022 | 0.9862 | 0.0047 | 0.8119 | 0.9560 |
| ***V119 (nodes 14 – 16) cEN – cEN*** | -0.0129 | 0.3403 | 0.9862 | -0.0043 | 0.8221 | 0.9560 |
| ***V172 (nodes 1-20) DMN – DMN / cEN*** | -0.0059 | 0.6100 | 0.9862 | 0.0268 | 0.1015 | 0.6769 |
| ***V174 (nodes 3-20) SN – DMN / CEN*** | -0.0108 | 0.4081 | 0.9862 | 0.0371 | 0.0446 | 0.6769 |
| ***V176 (nodes 5-20) cEN / DMN – DMN / cEN*** | -0.0018 | 0.8799 | 0.9862 | 0.0386 | 0.0247 | 0.6769 |
| ***V177 (nodes 6-20) cEN – DMN / cEN*** | -0.0033 | 0.7769 | 0.9862 | 0.0136 | 0.4157 | 0.8884 |
| ***V178 (nodes 7-20) DMN / cEN – DMN / cEN*** | 0.0113 | 0.3201 | 0.9862 | -0.0123 | 0.4469 | 0.8884 |
| ***V180 (nodes 9-20) cEN – DMN / cEN*** | 0.0082 | 0.5156 | 0.9862 | -0.0245 | 0.1708 | 0.6769 |
| ***V184 (nodes 13-20) DMN – DMN / cEN*** | 0.0022 | 0.8820 | 0.9862 | 0.0038 | 0.8528 | 0.9560 |
| ***V185 (nodes 14-20) cEN – DMN / cEN*** | 0.0151 | 0.2829 | 0.9862 | -0.0033 | 0.8691 | 0.9560 |
| ***V191 (nodes 1-21) DMN – cEN / DMN*** | -0.0129 | 0.3199 | 0.9862 | 0.0069 | 0.7079 | 0.9232 |
| ***V193 (nodes 3-21) SN – cEN / DMN*** | -0.0049 | 0.7185 | 0.9862 | 0.0073 | 0.7032 | 0.9232 |
| ***V195 (nodes 5-21) cEN / DMN – cEN / DMN*** | 0.0003 | 0.9816 | 0.9862 | -0.0124 | 0.4523 | 0.8884 |
| ***V196 (nodes 6-21) cEN – cEN / DMN*** | -0.0093 | 0.3439 | 0.9862 | 0.0275 | 0.0521 | 0.6769 |
| ***V197 (nodes 7-21) DMN / cEN – cEN / DMN*** | 0.0149 | 0.2505 | 0.9862 | -0.0158 | 0.3882 | 0.8884 |
| ***V199 (nodes 9-21) cEN – cEN / DMN*** | -0.0028 | 0.8252 | 0.9862 | -0.0242 | 0.1846 | 0.6769 |
| ***V203 (nodes 13-21) DMN – cEN / DMN*** | 0.0020 | 0.8854 | 0.9862 | -0.0288 | 0.1436 | 0.6769 |
| ***V204 (nodes 14-21) cEN – cEN / DMN*** | 0.0218 | 0.1560 | 0.9862 | -0.0483 | 0.0284 | 0.6769 |
| ***V206 (nodes 16-21) cEN – cEN / DMN*** | -0.0153 | 0.1979 | 0.9862 | 0.0144 | 0.3935 | 0.8884 |
| ***V210 (nodes 20-21) DMN / cEN – cEN / DMN*** | 0.0021 | 0.8757 | 0.9862 | 0.0271 | 0.1646 | 0.6769 |

#### STROBE Statement—checklist of items that should be included in reports of observational studies

|  | Item No. | Recommendation | Page  No. | Relevant text from manuscript |
| --- | --- | --- | --- | --- |
| **Title and abstract** | 1 | (*a*) Indicate the study’s design with a commonly used term in the title or the abstract | 1 |  |
|  |  | (*b*) Provide in the abstract an informative and balanced summary of what was done and what was found | 2 |  |
| Introduction | | | |  |
| Background/rationale | 2 | Explain the scientific background and rationale for the investigation being reported | 4 |  |
| Objectives | 3 | State specific objectives, including any prespecified hypotheses | 5 |  |
| Methods | | | |  |
| Study design | 4 | Present key elements of study design early in the paper | 5 |  |
| Setting | 5 | Describe the setting, locations, and relevant dates, including periods of recruitment, exposure, follow-up, and data collection | 5, SM |  |
| Participants | 6 | (*a*) *Cohort study*—Give the eligibility criteria, and the sources and methods of selection of participants. Describe methods of follow-up  *Case-control study*—Give the eligibility criteria, and the sources and methods of case ascertainment and control selection. Give the rationale for the choice of cases and controls  *Cross-sectional study*—Give the eligibility criteria, and the sources and methods of selection of participants | 5-8, SM |  |
|  |  | (*b*) *Cohort study*—For matched studies, give matching criteria and number of exposed and unexposed  *Case-control study*—For matched studies, give matching criteria and the number of controls per case | 6, SM |  |
| Variables | 7 | Clearly define all outcomes, exposures, predictors, potential confounders, and effect modifiers. Give diagnostic criteria, if applicable | 6-9, SM |  |
| Data sources/ measurement | 8* | For each variable of interest, give sources of data and details of methods of assessment (measurement). Describe comparability of assessment methods if there is more than one group | 6-9, SM |  |
| Bias | 9 | Describe any efforts to address potential sources of bias | 8-9, SM |  |
| Study size | 10 | Explain how the study size was arrived at | 5, SM |  |

Continued on next page

| Quantitative variables | 11 | Explain how quantitative variables were handled in the analyses. If applicable, describe which groupings were chosen and why | 9, SM |
| --- | --- | --- | --- |
| Statistical methods | 12 | (*a*) Describe all statistical methods, including those used to control for confounding | 9, SM |
|  |  | (*b*) Describe any methods used to examine subgroups and interactions | 9, SM |
|  |  | (*c*) Explain how missing data were addressed | 9-10, SM |
|  |  | (*d*) *Cohort study*—If applicable, explain how loss to follow-up was addressed  *Case-control study*—If applicable, explain how matching of cases and controls was addressed  *Cross-sectional study*—If applicable, describe analytical methods taking account of sampling strategy | NA |
|  |  | (*e*) Describe any sensitivity analyses | 9-10, SM |
| Results | | | |
| Participants | 13* | (a) Report numbers of individuals at each stage of study—eg numbers potentially eligible, examined for eligibility, confirmed eligible, included in the study, completing follow-up, and analysed | SM Fig 1 |
|  |  | (b) Give reasons for non-participation at each stage | SM Fig 1 |
|  |  | (c) Consider use of a flow diagram | SM Fig 1 |
| Descriptive data | 14* | (a) Give characteristics of study participants (eg demographic, clinical, social) and information on exposures and potential confounders | 10 |
|  |  | (b) Indicate number of participants with missing data for each variable of interest | 10, 13 |
|  |  | (c) *Cohort study*—Summarise follow-up time (eg, average and total amount) | 10 |
| Outcome data | 15* | *Cohort study*—Report numbers of outcome events or summary measures over time | 10-11 |
|  |  | *Case-control study—*Report numbers in each exposure category, or summary measures of exposure |  |
|  |  | *Cross-sectional study—*Report numbers of outcome events or summary measures |  |
| Main results | 16 | (*a*) Give unadjusted estimates and, if applicable, confounder-adjusted estimates and their precision (eg, 95% confidence interval). Make clear which confounders were adjusted for and why they were included | 10-14, SM |
|  |  | (*b*) Report category boundaries when continuous variables were categorized | 9, SM (confounds) |
|  |  | (*c*) If relevant, consider translating estimates of relative risk into absolute risk for a meaningful time period |  |

Continued on next page

| Other analyses | 17 | Report other analyses done—eg analyses of subgroups and interactions, and sensitivity analyses | 12-13 |
| --- | --- | --- | --- |
| Discussion | | | |
| Key results | 18 | Summarise key results with reference to study objectives | 14-15 |
| Limitations | 19 | Discuss limitations of the study, taking into account sources of potential bias or imprecision. Discuss both direction and magnitude of any potential bias | 17-18 |
| Interpretation | 20 | Give a cautious overall interpretation of results considering objectives, limitations, multiplicity of analyses, results from similar studies, and other relevant evidence | 15-17 |
| Generalisability | 21 | Discuss the generalisability (external validity) of the study results | 18 |
| Other information | |  | |
| Funding | 22 | Give the source of funding and the role of the funders for the present study and, if applicable, for the original study on which the present article is based | 19 |
